# Dynamical Effects of Homologous Reinfections in a Multi-Strain Dengue Model

**DOI:** 10.64898/2026.07.30.26359356

**Authors:** Akhil Kumar Srivastav, Vanessa Steindorf, Nico Stollenwerk, Bob W. Kooi, Maíra Aguiar

## Abstract

Dengue transmission is shaped by multiple viral serotypes, temporary cross-immunity (TCI), antibody-dependent enhancement (ADE), and repeated exposure in endemic populations. Classical multi-strain models usually assume lifelong protection against reinfection with the same serotype. However, recent evidence suggests that homologous dengue reinfections, although rare, can occur. Their population-level consequences remain poorly understood.

We extend a two-infection, two-strain dengue model with TCI and ADE-mediated transmission differences to include homologous reinfections. Homologous reinfection is represented by two exploratory parameters: relative susceptibility to reinfection with the same serotype and relative infectiousness during homologous reinfection. Using equilibrium analysis, bifurcation diagrams, simulations, and phase-space projections, we examine how these parameters affect dengue dynamics and interact with TCI duration and seasonal forcing under intermediate and long TCI durations, with and without seasonality.

The extended model shows that qualitative dynamics characteristic of endemic dengue transmission are reproduced mainly when susceptibility to homologous reinfection is low, so that homologous reinfections remain rare but dynamically influential. Longer TCI broadens regions of complex oscillatory dynamics, while seasonality shifts the bifurcation structure and makes torus bifurcations a central route to complex behavior. Although backward bifurcation can occur when homologous susceptibility exceeds the biologically meaningful range, this result should be interpreted as a mathematical mechanism rather than a realistic dengue scenario.

These results indicate that rare homologous reinfection pathways can influence long-term dengue dynamics when interacting with immune history, TCI, ADE-mediated transmission differences, and seasonal variation. Incorporating such pathways may improve understanding of recurrent outbreaks and irregular incidence patterns in highly exposed populations.

## 1 Introduction

Dengue fever, a viral disease transmitted primarily by *Aedes* mosquitoes, remains a major global public health concern, affecting millions of people annually across tropical and subtropical regions. Recent years have seen unprecedented dengue activity globally, followed by continued substantial transmission and expanding risk in both endemic and previously non-endemic regions. This expansion is particularly concerning where climatic suitability, human mobility, and vector establishment may facilitate local transmission. Alarmingly, autochthonous transmission has also been reported in regions previously considered non-endemic, including parts of Italy, France, and Spain [15]. Recent modeling work has further shown that dengue outbreaks in non-endemic areas may arise from stochastic introductions even under marginal transmission conditions, emphasizing the role of importation events in disease emergence [51, 22, 35, 52]. Since the beginning of 2026, and as of 23 March 2026, over 500 000 cases of dengue and over 100 dengue-related deaths had been reported globally, according to information from publicly available sources [16].

At the same time, several regions have experienced the re-emergence of viral serotypes that had not circulated locally for years, raising concerns about waning population immunity, changes in serotype dominance, and the potential for more severe outbreaks [38, 33]. These observations highlight the need for mathematical models that account for immune history, serotype interactions, reinfection pathways, and the possibility that epidemic emergence may be shaped by stochastic introductions as well as endemic transmission.

There are four dengue virus serotypes (DENV-1 to DENV-4). Infection with one serotype typically confers long-lasting protection against reinfection by the same serotype and short-term protection against the others, a phenomenon known as temporary cross-immunity (TCI). For decades, homologous reinfection, defined as reinfection with the same serotype, was considered highly unlikely because of durable serotype-specific immunity [21, 43, 57]. However, recent cohort studies have shown that such reinfections, although rare, can occur and may involve detectable viremia and measurable antibody responses [20, 46, 55, 40]. In a pediatric cohort study in Nicaragua, for example, 13.8% (4 out of 29) of homologous reinfections occurred between 325 and 621 days after the initial infection [55], and recent within-host models have reproduced patterns consistent with these observations [41, 7, 8]. These findings challenge the common modeling assumption that serotype-specific immunity is always lifelong and sterilizing. They also motivate the use of relative susceptibility and relative infectiousness parameters to represent, at the population level, incomplete serotype-specific protection and the potentially lower transmission contribution of homologous reinfections.

Most primary dengue infections are mild or asymptomatic, while heterologous secondary infections, defined as reinfections with a different serotype, are more often associated with severe outcomes such as dengue hemorrhagic fever and dengue shock syndrome, which can be fatal without timely treatment [10, 16, 21, 45]. This increased severity is commonly linked to antibody-dependent enhancement (ADE), whereby non-neutralizing or sub-neutralizing antibodies from a prior infection facilitate viral entry into host cells, increasing viral replication and worsening symptoms [23, 24, 26, 27, 39, 42]. ADE may also affect transmission dynamics, since severe cases may experience reduced mobility or hospitalization and may therefore contribute differently to onward transmission compared with mild or asymptomatic infections. These immune-mediated mechanisms are also relevant for dengue vaccination, since vaccine performance and safety depend on prior exposure, serostatus, and the ability to induce balanced protection across serotypes. Thus, understanding how serotype-specific protection, cross-immunity, and reinfection shape population-level dynamics remains important not only for interpreting natural epidemics, but also for anticipating the epidemiological consequences of immune interventions.

Mathematical modeling of dengue dates back to the early 1970s [25], and has since evolved to incorporate the interplay between serotype dynamics, TCI, and ADE [2, 47, 48, 9]. These models have shown that dengue transmission can exhibit oscillatory and chaotic dynamics, particularly when multi-strain interactions and immune-mediated mechanisms are included [1, 5, 4, 29, 17, 18, 44]. Such dynamics have been used to interpret irregular fluctuations in dengue incidence, including patterns observed in Thailand [3, 4]. Together, this body of work supports the broader modelling rationale for relaxing restrictive immunity assumptions and for examining whether rare reinfection pathways can affect nonlinear dengue dynamics at the population level.

Despite these advances, most multi-strain dengue models exclude homologous reinfections by assuming complete lifelong protection against the infecting serotype. This assumption may be overly restrictive, particularly when repeated viral exposure, immune boosting, and waning protection interact over long time scales. Even if homologous reinfections are rare at the individual level, they may alter population-level dynamics by modifying the pool of infectious individuals, changing outbreak timing, and shifting the stability of endemic states.

Here, we investigate the dynamical consequences of relaxing the classical assumption of complete homologous protection. We refine and extend the multi-strain dengue model from [3] by explicitly incorporating reinfections with the same serotype alongside primary and heterologous secondary infections. The extended model includes TCI, ADE-mediated changes in transmission during heterologous secondary infections, and two additional parameters: relative susceptibility to reinfection with the same serotype and relative infectiousness during such reinfections. These parameters are treated as exploratory sensitivity parameters, reflecting the current uncertainty surrounding the frequency, infectiousness, and epidemiological contribution of homologous dengue reinfections. Using numerical bifurcation analysis and time-series simulations, we characterize how these reinfection pathways, together with TCI and ADE-mediated changes in transmission, alter the qualitative dynamics of dengue in highly exposed populations, including transitions among stable endemic persistence, periodic outbreaks, quasi-periodic behavior, and chaotic epidemic fluctuations.

## 2 Mathematical model

The two-infection multi-strain dengue model proposed in [1, 3] demonstrated that dengue transmission can exhibit chaotic dynamics when immunological features such as TCI and ADE are incorporated. In particular, when ADE-associated severe cases are assumed to contribute less to the force of infection, the dynamics become even more complex for intermediate durations of TCI. Subsequent work showed that these dynamics are driven primarily by the interaction between TCI and ADE-mediated differences between primary and secondary infections, rather than by the detailed number of strains explicitly represented [4]. Bifurcation analysis of a simplified two-infection SIR-SIR model further showed that temporary immunity and disease enhancement can shape rich stationary and oscillatory dynamics, highlighting TCI duration as a key dynamical parameter [6]. Motivated by these results and by the evidence discussed above, we relax the classical assumption of complete lifelong homologous protection and extend the baseline framework to include reinfection with the same serotype.

The resulting compartmental model, illustrated in Fig. 1, divides the total population *N* into 12 classes: susceptible naive, or seronegative, individuals (*S*), individuals undergoing a primary infection with strain one or strain two (*I*_1_, *I*_2_), recovered individuals with temporary cross-protection (*R*_1_, *R*_2_), seropositive individuals who have recovered from a primary infection with strain one or strain two and have lost TCI (*S*_1_, *S*_2_), individuals with heterologous secondary infections (*I*_21_, *I*_12_), individuals experiencing homologous reinfection with strain one or strain two (*I*_11_, *I*_22_), and fully recovered individuals (*R*). Throughout the model, *I*_*ij*_ denotes individuals previously infected with strain *i* who are currently infected with strain *j*. Thus, *I*_12_ and *I*_21_ represent heterologous secondary infections, whereas *I*_11_ and *I*_22_ represent homologous reinfections.

**Figure 1:**
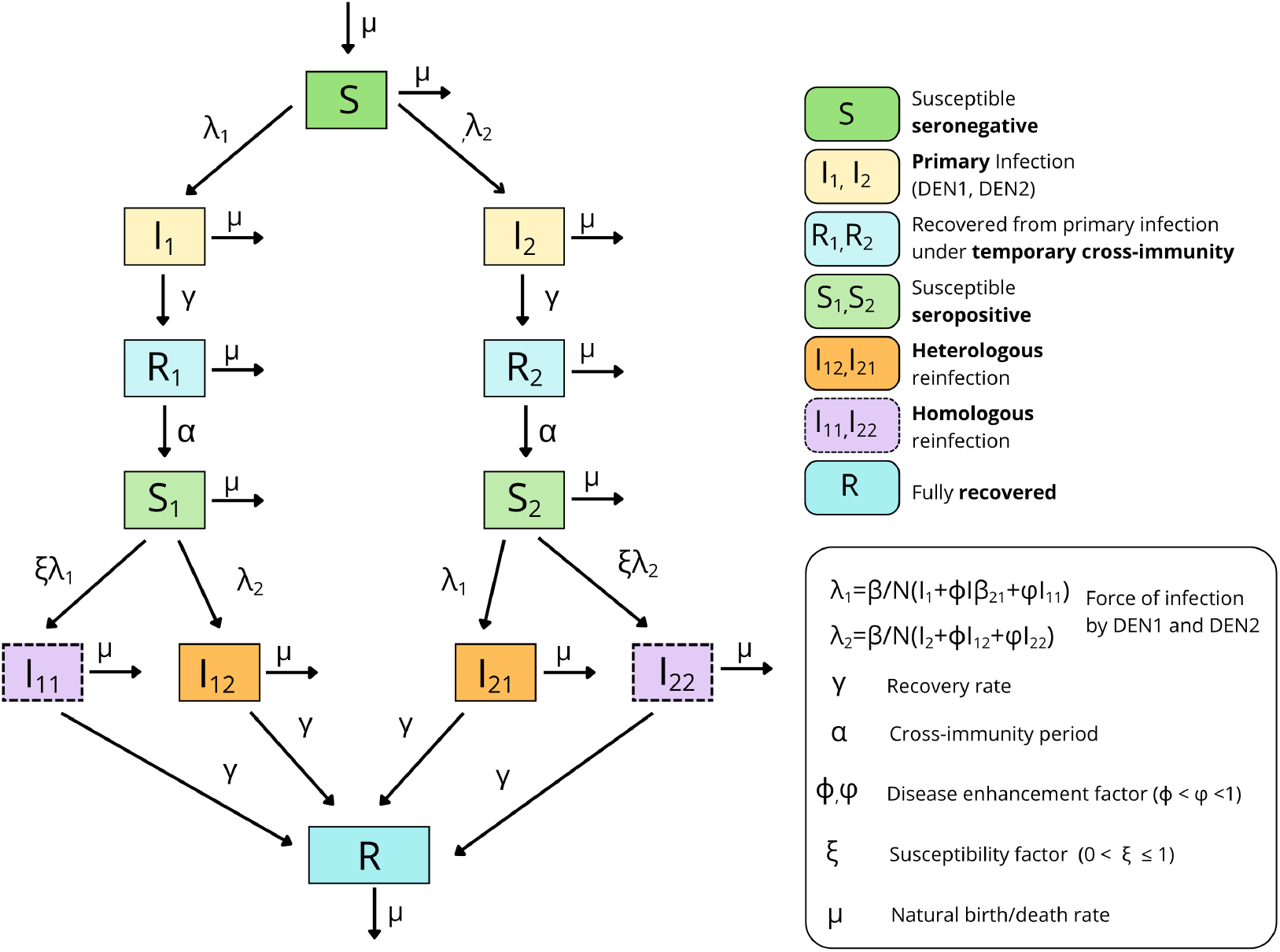
Flow chart of the extended two-strain dengue compartmental model, including homologous reinfections. The quantities *λ*_1_ and *λ*_2_ denote the forces of infection generated by individuals currently infected with strains one and two, respectively. The parameter *ϕ* denotes the relative infectiousness of heterologous secondary infections, *φ* denotes the relative infectiousness of homologous reinfections, and homologous susceptibility is scaled by *ξ*.

The force of infection generated by individuals currently infected with strain one or strain two is given by

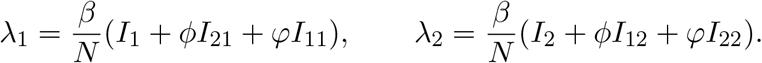

Thus, seronegative individuals acquire primary infections with strain one or strain two at rates *λ*_1_ and *λ*_2_, respectively. Seropositive individuals in *S*_1_ may acquire a heterologous secondary infection with strain two at rate *λ*_2_ or a homologous reinfection with strain one at rate *ξλ*_1_. Similarly, individuals in *S*_2_ may acquire a heterologous secondary infection with strain one at rate *λ*_1_ or a homologous reinfection with strain two at rate *ξλ*_2_. Therefore, the total forces acting on *S*_1_ and *S*_2_ are

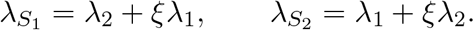

Here, *β* is the baseline transmission rate, *ϕ* denotes the relative infectiousness of heterologous secondary infections, *φ* denotes the relative infectiousness of homologous reinfections, and *ξ* represents reduced susceptibility to homologous reinfection due to partial serotype-specific immunity. We assume *ϕ < φ <* 1, reflecting the lower contribution of severe heterologous secondary infections to transmission and the higher mobility, but lower viremia, expected during homologous reinfections [3, 4, 8, 39]. The parameters *ξ* and *φ* are introduced as exploratory biological parameters rather than as directly fitted quantities. The parameter *ξ* captures residual susceptibility to homologous reinfection after previous exposure to the same serotype, with *ξ* = 0 corresponding to complete sterilizing homologous protection and 0 *< ξ <* 1 representing incomplete or partially waned protection. Because homologous reinfections are expected to be rare, low values of *ξ* are the most biologically plausible, whereas values close to one should be interpreted as upper-bound sensitivity scenarios. The parameter *φ* captures the relative infectiousness of homologously reinfected individuals compared with primary infections. Pre-existing serotype-specific immunity may reduce viral replication or shorten viremia, while milder disease may also imply less reduction in mobility than during severe heterologous secondary infections. For this reason, we consider 0 ≤ *φ* ≤ 1 and use *ϕ < φ <* 1 as a biologically motivated baseline assumption.

The transmission dynamics occur within a constant host population size *N* . Vector dynamics are represented implicitly through effective transmission parameters. This reduction follows previous work showing that, for the type of epidemic dynamics considered here, explicit mosquito compartments may not qualitatively change the main transmission patterns [50].

After a primary infection, individuals recover at rate *γ* and enter a period of TCI, during which high levels of cross-reactive antibodies provide protection against all serotypes. Loss of cross-immunity occurs at rate *α*, so that the average duration of TCI is 1*/α*. Literature estimates typically place this period between 3 and 9 months [37], though some studies suggest that it may last up to 2 years [32, 28]. To investigate the impact of TCI duration on transmission dynamics, our simulations consider two average durations: intermediate, with 1*/α* = 6 months, corresponding to *α* = 2, year^−1^, and long, with 1*/α* = 2 years, corresponding to *α* = 0.5, year^−1^.

Supported by cohort and immunological studies [12, 34], as well as previous modeling work [3, 56, 53, 31], the model considers at most two infections per individual, regardless of their order or type. This is based on evidence that subsequent clinical infections, such as tertiary or quaternary infections, are rarely reported. Moreover, since most dengue infections are asymptomatic, it remains unclear whether and to what extent additional reinfections contribute meaningfully to transmission dynamics. After two infections, individuals are assumed to acquire full protection against all serotypes and transition to the fully recovered class *R*.

The model is based on the following assumptions: (i) the host population size is constant, with births and deaths occurring at rate *µ*; (ii) the two dengue strains are epidemiologically symmetric; (iii) vector dynamics are represented implicitly through effective transmission parameters; (iv) individuals experience at most two infections; (v) homologous reinfections occur with reduced susceptibility, controlled by *ξ*; (vi) the infectious contribution of homologous reinfections is controlled by *φ* and is explored as a sensitivity parameter; and (vii) heterologous secondary infections may contribute less to transmission than primary infections because severe ADE-associated cases are more likely to experience reduced mobility or hospitalization.

All state variables are non-negative. Unless otherwise stated, the biologically meaningful parameter ranges are *β, γ, α, µ >* 0, 0 ≤ *ϕ* ≤ 1, 0 ≤ *φ* ≤ 1, and 0 ≤ *ξ* ≤ 1. Values outside these ranges are used only, when explicitly stated, for mathematical continuation and not as biologically realistic dengue scenarios.

The total host population is given by

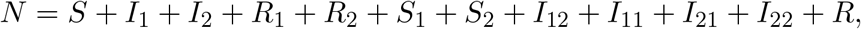

and satisfies 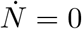.

The system of ordinary differential equations governing the dynamics of dengue transmission, including primary infections, heterologous secondary infections, and homologous reinfections, is expressed as

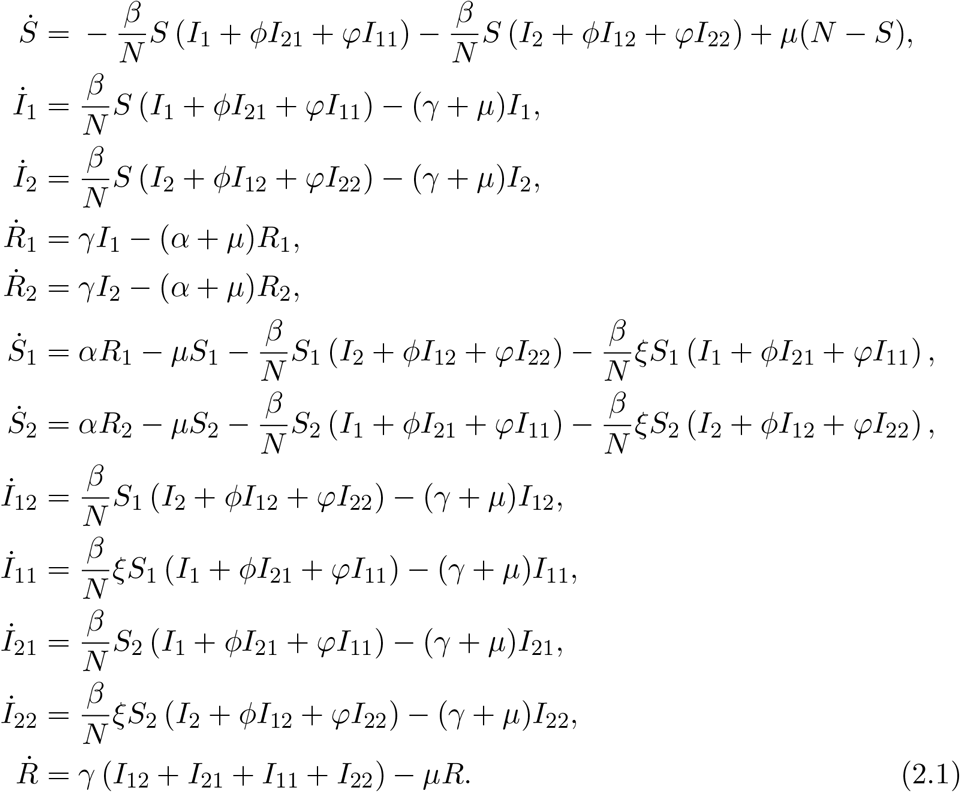

For simplicity, and consistent with previous analyses showing that large strain asymmetries can undermine coexistence and lead to the extinction of one strain, we assume no epidemiological asymmetry between the two strains [29]. Thus, infections with strain one or strain two contribute equally to the force of infection.

## 3 Mathematical Analysis

### 3.1 Positivity and boundedness of the extended model

We first show that the solutions of system (2.1) remain non-negative and bounded for non-negative initial conditions. Evaluating the vector field on the boundary of the non-negative state space gives

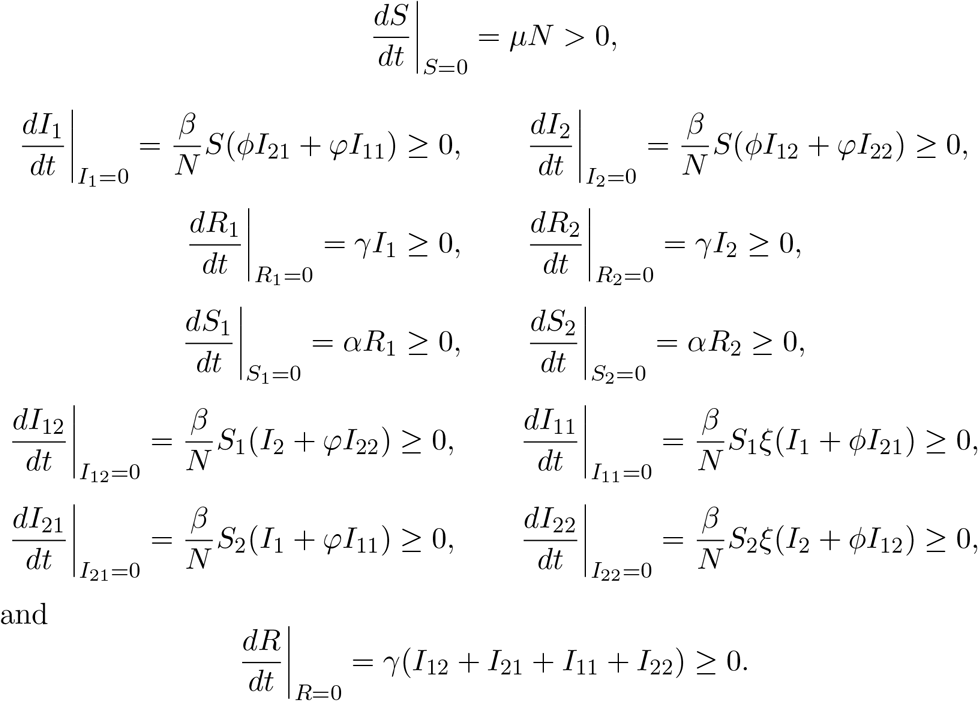

Thus, the vector field points inward or is tangent to the boundary of the non-negative orthant. Therefore, for non-negative initial conditions, all state variables remain non-negative for all *t* ≥ 0.

Moreover, summing all equations in system (2.1) gives

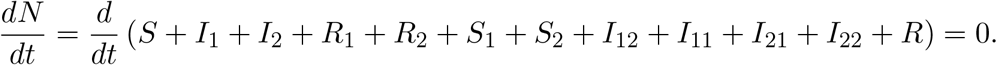

Hence, the total host population remains constant. Since all state variables are non-negative and their sum is *N*, each compartment is bounded above by *N* . Therefore, the biologically feasible region is

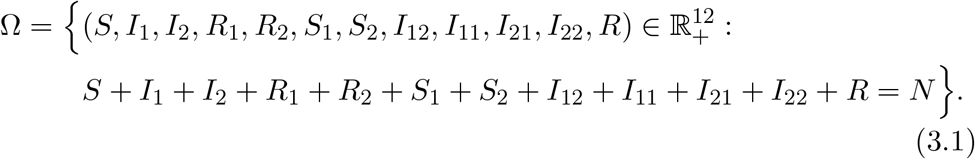

Thus, the following result holds.

#### Theorem 1.

*The biologically feasible region* Ω *is positively invariant under system (2*.*1)*.

### 3.2 The Disease-Free Equilibrium (DFE) and the reproduction number ℛ_0_

In this section, we establish the disease-free equilibrium (DFE) and compute the basic reproduction number ℛ_0_ using the next-generation matrix method. The reproduction number gives the average number of secondary infections generated by a typical infectious individual introduced into an entirely susceptible population.

#### Theorem 2.

*The system (2*.*1) has a disease-free equilibrium in* Ω *given by*

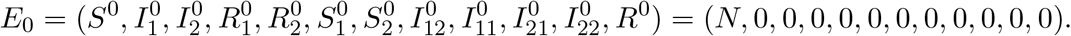

*Proof*. At the disease-free state, all infected compartments are zero. Since there is no prior exposure in the population, the recovered and seropositive compartments are also zero. Hence, the entire population is in the susceptible class, giving

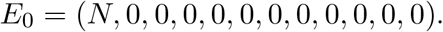

Substitution into system (2.1) verifies that all right-hand sides vanish at this point.

To compute ℛ_0_, we consider the infected compartments

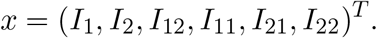

Let ℱ denote the vector of new infection terms and *V* the vector of transition terms out of infected compartments. These are given by

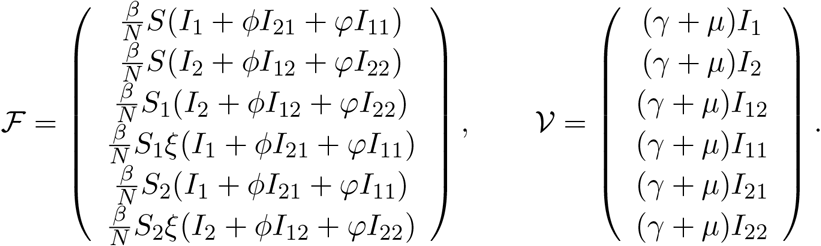

Evaluating the Jacobian matrices of ℱ and *V* at the DFE gives

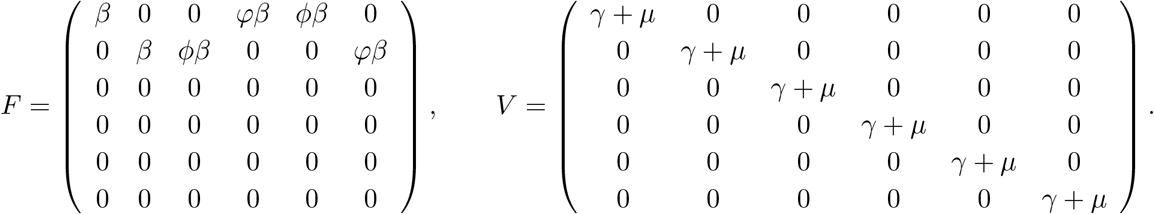

The next-generation matrix is *K* = *FV* ^−1^, and the basic reproduction number is defined as its spectral radius [54]:

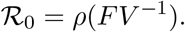

For this system,

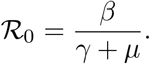

This expression reflects that, at the DFE, the population is entirely seronegative; hence, only the primary infection compartments are epidemiologically accessible at invasion. The homologous and heterologous reinfection pathways do not contribute to invasion at the DFE, although they can strongly affect endemic and oscillatory dynamics once infection has circulated in the population.

Thus, ℛ_0_ governs local invasion near the disease-free equilibrium, but it does not by itself characterize the richer nonlinear dynamics generated by TCI, ADE, and homologous reinfection away from the DFE. This distinction is important because the homologous reinfection parameters *ξ* and *φ* do not affect invasion into a fully susceptible population, but they can affect endemic and post-invasion dynamics once immune history has accumulated in the population.

### 3.3 The Disease Endemic Equilibrium (DEE)

We next investigate the existence of endemic equilibria for system (2.1). At equilibrium, the symmetry of the two-strain formulation allows the force-of-infection variables to be reduced to algebraic relations involving *λ*_1_ and *λ*_2_. Solving the equilibrium equations yields two possible branches: a symmetric biologically meaningful branch with *λ*_1_ = *λ*_2_, and a second algebraic branch with *λ*_2_ = −(*λ*_1_ + *µ*), which is biologically inadmissible because forces of infection must be non-negative.

**Case 1:** *λ*_2_ = *λ*_1_

For the symmetric endemic branch, substituting *λ*_2_ = *λ*_1_ into the equilibrium relations yields the quadratic equation

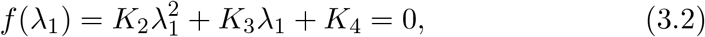

where

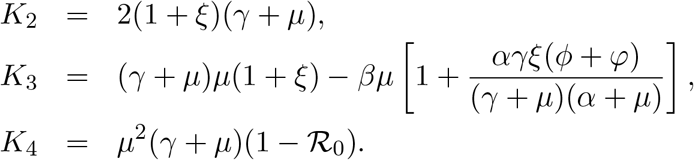

Here, *K*_2_ *>* 0, while the sign of *K*_3_ depends on *ξ* and the remaining epidemiological parameters. The sign of *K*_4_ is determined by ℛ_0_. The possible sign combinations and the corresponding number of positive roots, obtained using Descartes’ rule of signs, are summarized in Table 1.

**Table 1:** Descartes’ rule of signs applied to the polynomial *f* (*λ*_1_) in equation (3.2). When ℛ_0_ *>* 1, the polynomial has exactly one positive root. When ℛ_0_ *<* 1, two positive roots may occur only if *K*_3_ *<* 0.

| if $\mathcal{R}_0 > 1$ | $K_2$ | $K_3$ | $K_4$ | Changes of signs | Positive roots |
| --- | --- | --- | --- | --- | --- |
| i) | + | + | - | 1 | one |
| ii) | + | - | - | 1 | one |
| if $\mathcal{R}_0 < 1$ | $K_2$ | $K_3$ | $K_4$ | Changes of signs | Positive roots |
| i) | + | + | + | 0 | none |
| ii) | + | - | + | 2 | two or none |

Since *K*_2_ *>* 0, the number of positive roots depends on the signs of *K*_3_ and *K*_4_. If *R*_0_ *>* 1, then *K*_4_ *<* 0, and the polynomial has exactly one positive root, independently of the sign of *K*_3_. Therefore, the system admits a unique positive endemic equilibrium on the symmetric branch.

If ℛ_0_ *<* 1, then *K*_4_ *>* 0. In this case, no positive endemic equilibrium exists when *K*_3_ *>* 0. When *K*_3_ *<* 0, Descartes’ rule allows either two positive roots or no positive roots; two positive roots occur only when the discriminant is positive and both roots are biologically admissible. This case corresponds to the mathematical possibility of backward bifurcation and bistability, where a locally stable disease-free equilibrium may coexist with endemic equilibria.

The condition *K*_3_ *<* 0 can be written as

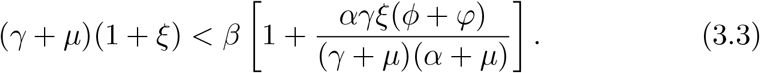

This inequality identifies a necessary condition under which the polynomial may admit two positive endemic-equilibrium roots when ℛ_0_ *<* 1. The actual existence of two biologically meaningful equilibria also requires a positive discriminant and positive admissible roots.

Figure 2 illustrates the two cases. Panel (a) shows a biologically meaningful value of *ξ*, for which a unique endemic equilibrium appears when ℛ_0_ *>* 1. Panel (b) shows a backward-bifurcation structure obtained for *ξ* = 4.

**Figure 2:**
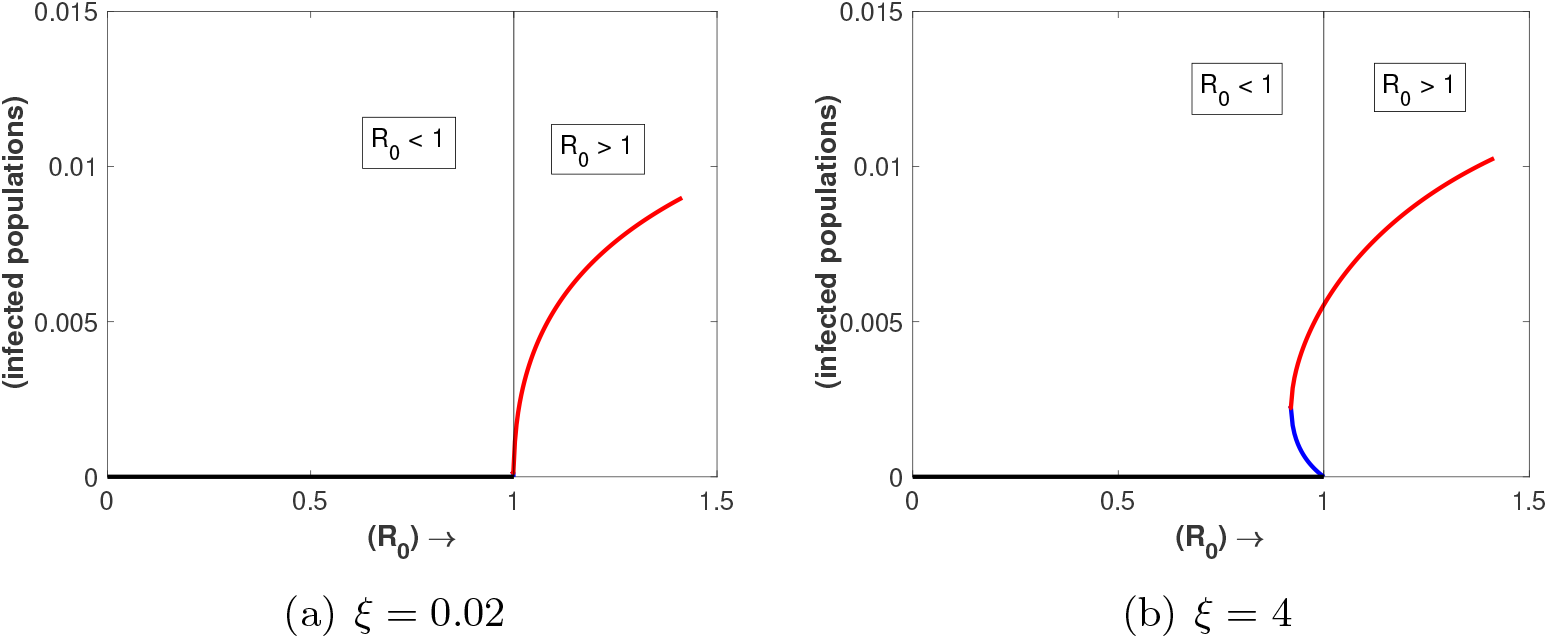
Steady states for the total infected population, *I* = *I*_1_ + *I*_2_ + *I*_11_ + *I*_12_ + *I*_21_ + *I*_22_, as the transmission parameter *β* varies. Panel (a) shows a biologically meaningful case with *ξ* = 0.02. Panel (b) shows *ξ* = 4, which lies outside the biologically meaningful range 0 ≤ *ξ* ≤ 1 and is included only to illustrate the mathematical possibility of bistability and backward bifurcation. The remaining parameters are fixed as in Table 2.

Because *ξ* represents relative susceptibility to homologous reinfection, biologically meaningful values satisfy 0 ≤ *ξ* ≤ 1. Therefore, the bistability and backward bifurcation illustrated for *ξ* = 4 should be interpreted only as a mathematical continuation result, not as a biologically realistic dengue scenario. This case shows that the model structure is capable of producing multiple endemic equilibria when the contribution of homologous reinfection is artificially amplified. For the biologically relevant range considered in the numerical simulations, 0 ≤ *ξ* ≤ 1, the backward-bifurcation branch is not expected under the baseline parameterization. Accordingly, the backward-bifurcation result is retained to document the mathematical structure of the extended model, whereas the epidemiological interpretation of the results focuses on the biologically meaningful range of homologous susceptibility.

**Table 2:** Baseline parameter values used for simulations. Values are based on [1, 3], with homologous reinfection parameters introduced in the extended model. The values of *ξ* and *φ* are not fitted estimates, but baseline values used for numerical exploration and sensitivity analysis.

| Parameter | Description | Baseline value |
| --- | --- | --- |
| $\beta$ | effective transmission rate | $2\gamma$ |
| $\gamma$ | recovery rate | $52 \text{ year}^{-1}$ (1 week) |
| $\alpha$ | TCI waning rate | $0.5 \text{ year}^{-1}$ (2 years)<br>or $2 \text{ year}^{-1}$ (6 months) |
| $\phi$ | relative infectiousness<br>of heterologous secondary infections | 0.8 |
| $\mu$ | natural birth and mortality rate | $1/65 \text{ year}^{-1}$ |
| $\varphi$ | relative infectiousness of homologous reinfections | 0.95 |
| $\xi$ | relative susceptibility to homologous reinfection | 0.9 |
| $\eta$ | seasonal forcing amplitude | 0 or 0.35 |
| $T$ | seasonal period | 1 year |

**Case 2:** *λ*_2_ = −(*λ*_1_ + *µ*)

The second algebraic branch is biologically inadmissible. Forces of infection must satisfy *λ*_1_ ≥ 0 and *λ*_2_ ≥ 0. However, if *λ*_2_ = −(*λ*_1_ +*µ*) with *λ*_1_ ≥ 0 and *µ >* 0, then *λ*_2_ *<* 0, which is impossible in the biologically feasible region. Therefore, no endemic equilibrium exists on this branch.

These results are summarized in the following theorem.

#### Theorem 3.

*For the symmetric endemic branch λ*_1_ = *λ*_2_, *if* ℛ_0_ *>* 1, *system (2*.*1) admits a unique positive endemic equilibrium. If* ℛ_0_ *<* 1, *then no positive endemic equilibrium exists when K*_3_ *>* 0, *whereas two positive endemic equilibria may exist when K*_3_ *<* 0 *and the roots of equation (3.2) are positive and biologically admissible. The algebraic branch λ*_2_ = −(*λ*_1_ + *µ*) *is biologically inadmissible*.

#### Remark 1.

*The case with* ℛ_0_ *<* 1, *K*_3_ *<* 0, *and two biologically admissible positive roots corresponds to a mathematical backward-bifurcation and bistability mechanism. However, under the baseline parameterization considered here, this mechanism requires values of ξ outside the biologically meaningful range. It should therefore be interpreted as an algebraic/dynamical possibility of the model rather than as a realistic dengue scenario*.

### 3.4 Stability analysis of the equilibria

In this section, we analyze the stability of the disease-free equilibrium and discuss the local stability of endemic equilibria. The stability of the DFE can be obtained analytically, while the stability of endemic equilibria is assessed through the eigenvalues of the Jacobian matrix evaluated at the corresponding equilibrium points.

#### Theorem 4.

*The disease-free equilibrium E*_0_ *of system (2*.*1) is locally asymptotically stable if* ℛ_0_ *<* 1 *and unstable if* ℛ_0_ *>* 1.

*Proof*. The disease-free equilibrium is

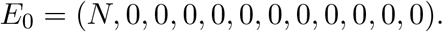

At this equilibrium, the only epidemiologically accessible infected compartments are the primary infection compartments *I*_1_ and *I*_2_, since the seropositive compartments are zero. Linearizing the infected subsystem around *E*_0_ gives the eigenvalues

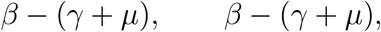

associated with the invasion of strains one and two. The remaining infected compartments, *I*_12_, *I*_11_, *I*_21_, and *I*_22_, have eigenvalues

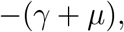

because no seropositive individuals are present at the DFE.

The non-infected compartments contribute eigenvalues

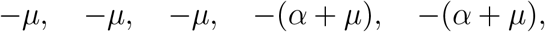

all of which are negative. Hence, all eigenvalues of the Jacobian at *E*_0_ have negative real parts if

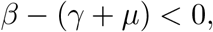

or equivalently,

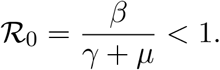

If ℛ_0_ *>* 1, then *β* − (*γ* + *µ*) *>* 0, and at least one eigenvalue is positive.

Therefore, *E*_0_ is locally asymptotically stable for ℛ_0_ *<* 1 and unstable for ℛ_0_ *>* 1.

#### Theorem 5.

*Assume that the parameters are in the biologically meaningful range*

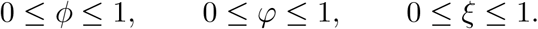

*If* ℛ_0_ *<* 1, *then the disease-free equilibrium E*_0_ *is globally asymptotically stable in the biologically feasible region* Ω.

*Proof*. Let

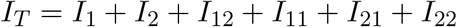

denote the total infected population. Summing the equations for all infected compartments gives

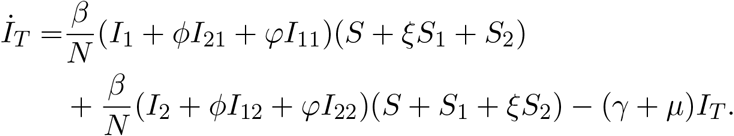

Since 0 ≤ *ϕ, φ, ξ* ≤ 1 and all state variables are non-negative, we have

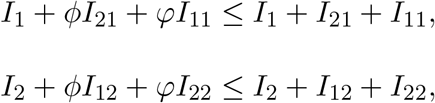

and

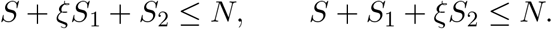

Therefore,

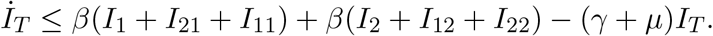

Thus,

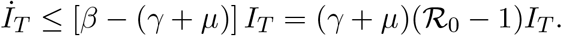

If ℛ_0_ *<* 1, then *İ*_*T*_ ≤ −*cI*_*T*_ for some *c >* 0, and hence

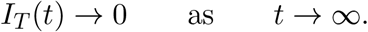

Consequently,

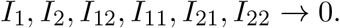

The equations for *R*_1_ and *R*_2_ are

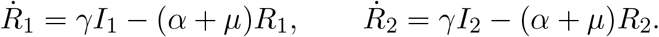

Since *I*_1_, *I*_2_ → 0, it follows that

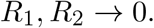

Similarly, the equations for *S*_1_ and *S*_2_ imply

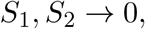

and the equation for *R* implies

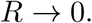

Because the total population is constant and equal to *N*, we obtain

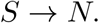

Therefore,

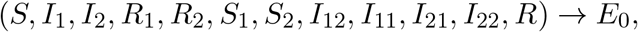

which proves that the disease-free equilibrium is globally asymptotically stable in Ω when ℛ_0_ *<* 1.

#### Remark 2.

The global stability result above is stated for the biologically meaningful parameter range 0 ≤ *ϕ, φ, ξ* ≤ 1. If parameters outside this range are used for mathematical continuation, for example *ξ >* 1 to illustrate backward bifurcation, the above comparison argument no longer applies directly.

Because the endemic equilibrium is high-dimensional and depends on multiple immune-history compartments, we combine analytical existence conditions with numerical continuation to characterize stability changes and bifurcation structure.

#### Local stability of endemic equilibria

Let

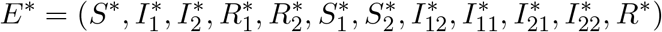

denote a positive endemic equilibrium of system (2.1). The local stability of *E*^∗^ is determined by the eigenvalues of the Jacobian matrix

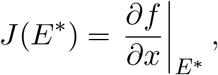

where

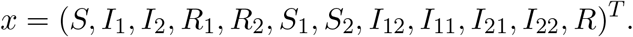

The endemic equilibrium is locally asymptotically stable if all eigenvalues of *J*(*E*^∗^) have negative real parts, and unstable if at least one eigenvalue has positive real part.

Equivalently, local asymptotic stability can be assessed from the characteristic polynomial

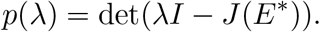

If this polynomial satisfies the Routh–Hurwitz conditions, then *E*^∗^ is locally asymptotically stable. Because the full system is 12-dimensional, the explicit Routh–Hurwitz determinants are algebraically intricate and provide limited interpretability. Therefore, in the numerical bifurcation analysis below, the stability of endemic equilibria is determined by numerical continuation and by monitoring the eigenvalues of the Jacobian along equilibrium branches.

## 4 Results

To investigate the impact of homologous reinfection under biologically plausible durations of TCI, we analyze the system under intermediate and long average periods of cross-protection. We first examine the autonomous baseline model using bifurcation diagrams, which identify parameter values at which qualitative changes in the long-term dynamics occur, including transitions between stable endemic equilibria, periodic oscillations, quasi-periodic behavior, and irregular dynamics consistent with chaos. We then complement these results with time-series simulations and phase-space projections to illustrate the corresponding epidemic trajectories.

We next introduce seasonal forcing in the transmission rate, allowing us to assess how periodic environmental variation reshapes the bifurcation structure and the routes to complex dynamics.

Numerical simulations were conducted in MATLAB [30], while bifurcation analysis was performed using AUTO [14, 36], a continuation software package for the analysis of ordinary differential equations. Table 2 summarizes the model parameters and baseline values used in the numerical experiments. The homologous reinfection parameters *ξ* and *φ* are treated as exploratory sensitivity parameters, while *α* and *η* are varied to assess the effects of TCI duration and seasonal forcing.

### 4.1 Numerical bifurcation analysis of the non-seasonal system

#### 4.1.1 One-parameter bifurcation diagrams

We first analyze the non-seasonal system by varying parameters associated with homologous reinfection. Previous bifurcation studies of two-infection dengue models showed that the interaction between TCI and ADE-mediated differences between primary and secondary infections can generate stable endemic equilibria, periodic oscillations, quasi-periodic dynamics, and chaotic or chaos-like behavior [3, 4, 6]. Building on this framework, we fix the baseline parameter values listed in Table 2 and investigate how the homologous susceptibility parameter *ξ* and the homologous transmissibility parameter *φ* affect the dynamics for two average durations of TCI: an intermediate duration of six months, corresponding to *α* = 2, year^−1^, and a longer duration of two years, corresponding to *α* = 0.5, year^−1^. This analysis provides a structured sensitivity assessment of the two parameters introduced to represent homologous reinfection, while also showing how their dynamical effects depend on TCI duration.

In all bifurcation diagrams, the total infected population is defined as

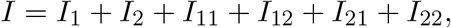

which includes primary infections, heterologous secondary infections, and homologous reinfections.

Figure 3 shows that the homologous susceptibility parameter *ξ* strongly modulates the qualitative dynamics of the system. For the intermediate TCI duration, *α* = 2, panels (a,b) indicate a region of irregular dynamics consistent with chaos for small values of *ξ*, approximately *ξ* ∈ (0, 0.15). In this region, epidemic trajectories are highly sensitive to initial conditions and display irregular fluctuations. As *ξ* increases, the system transitions from irregular dynamics to periodic behavior through bifurcations of limit cycles. Over intermediate values of *ξ*, stable periodic orbits are observed, corresponding to recurrent epidemic cycles. For larger values, approximately *ξ* ∈ (0.7, 1), the system approaches a stable endemic equilibrium, indicating persistence at an approximately constant level.

**Figure 3:**
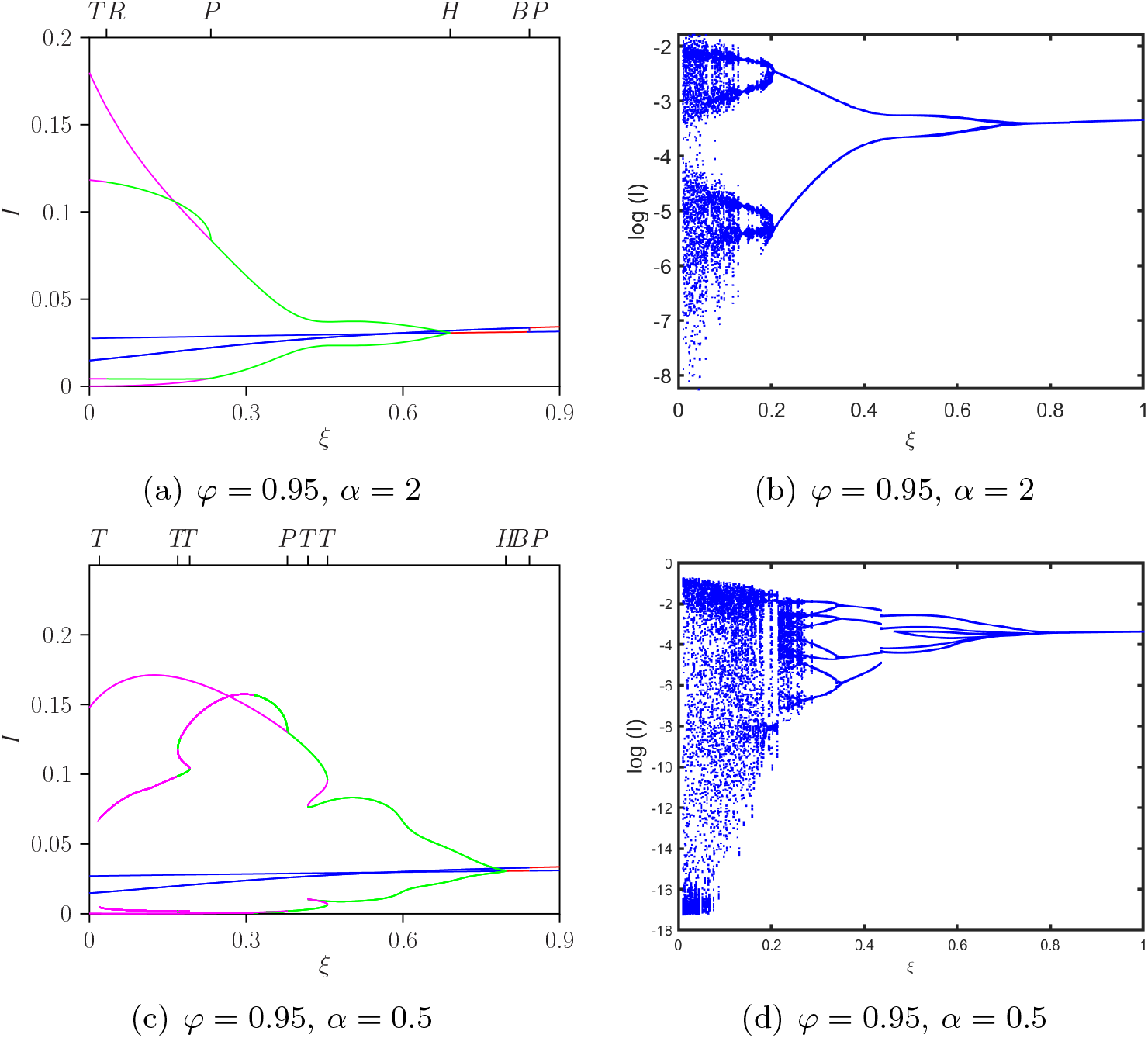
One-parameter bifurcation diagrams obtained by varying the homologous susceptibility parameter *ξ*, with *φ* = 0.95. Panels (a,b) correspond to 1*/α* = 6 months (*α* = 2, year^−1^), and panels (c,d) correspond to 1*/α* = 2 years (*α* = 0.5, year^−1^). Panels (a,c) show solution branches for the total infected population *I*, while panels (b,d) show corresponding extrema of ln(*I*). Labels indicate Hopf (*H*), pitchfork (*P* ), torus (*TR*), tangent/fold (*T* ), and boundary point (*BP* ) bifurcations, when present. The remaining parameters are fixed as in Table 2.

For the longer TCI duration, *α* = 0.5, panels (c,d) show a broader region of complex dynamics. A region of irregular dynamics consistent with chaos is again observed for small values of *ξ*, approximately *ξ* ∈ (0, 0.18), followed by intervals of periodic and more complex oscillatory behavior, including tangent and pitchfork bifurcations. For larger values of *ξ*, approximately *ξ* ∈ (0.75, 1), the system again converges to a stable endemic equilibrium. Thus, longer TCI increases the range of parameter values over which complex oscillatory dynamics can occur.

Varying *ξ* reveals that the qualitative dynamics typically associated with endemic dengue transmission, including recurrent outbreaks, multi-periodic oscillations, and irregular dynamics consistent with chaos, are reproduced when susceptibility to homologous reinfection remains low. As *ξ* increases, the system progressively loses these complex oscillatory regimes and approaches stable endemic persistence. Thus, homologous reinfection does not need to be frequent to influence dengue dynamics; rather, rare homologous reinfection pathways, combined with TCI and ADE-mediated differences in transmission, are sufficient to shape the long-term behavior of the system.

Figure 4 shows the corresponding dynamics when the homologous transmissibility parameter *φ* is varied while fixing *ξ* = 0.9. For the intermediate TCI duration, the qualitative pattern is broadly similar to that obtained when varying *ξ*: low values of *φ* can be associated with irregular dynamics consistent with chaos, intermediate values with periodic oscillations, and larger values with stable endemic persistence. However, because *φ* affects the infectious contribution of homologous reinfections rather than the susceptibility of seropositive individuals, the location and width of the irregular and periodic windows differ from those observed when varying *ξ*.

**Figure 4:**
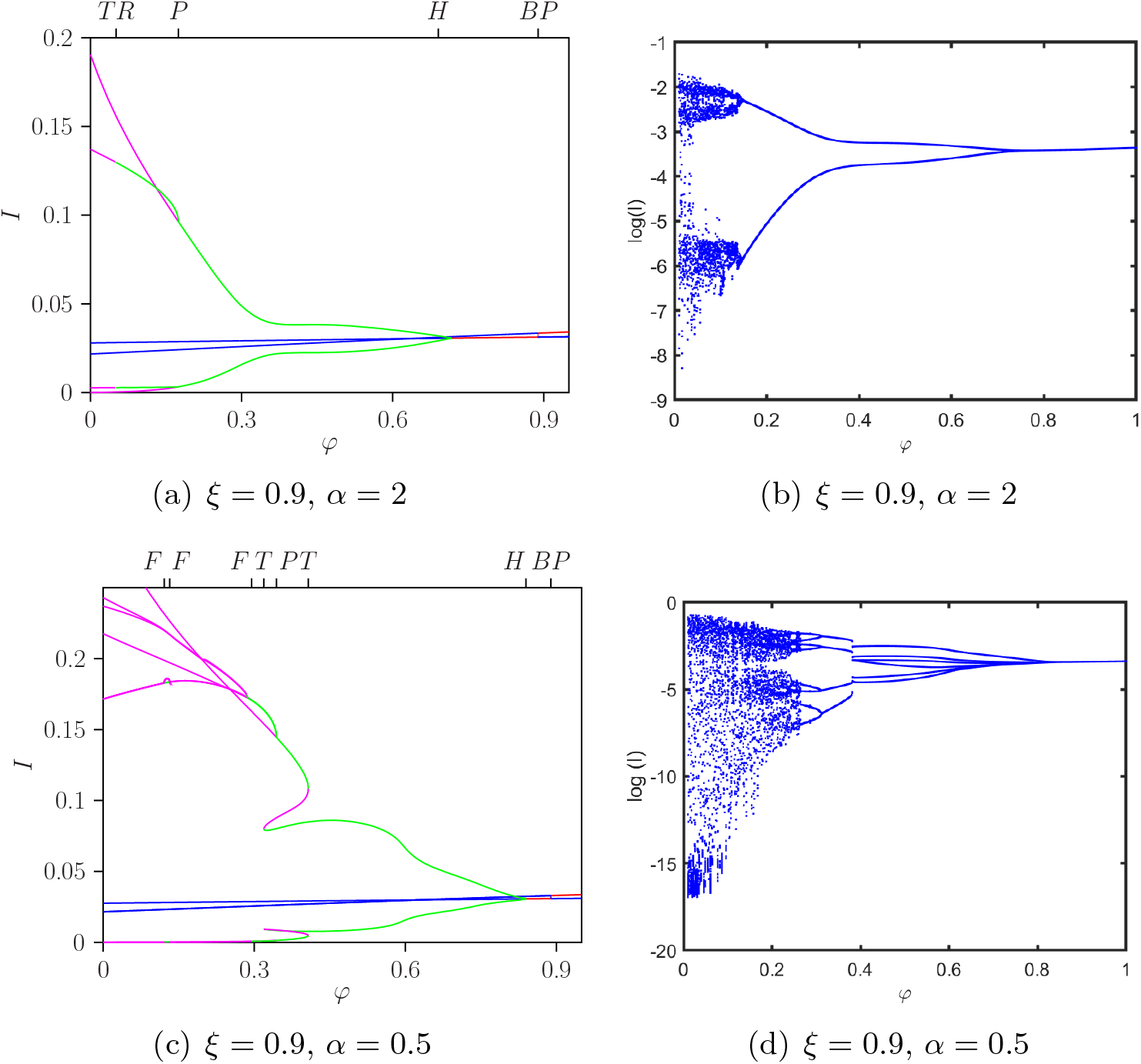
One-parameter bifurcation diagrams obtained by varying the homologous transmissibility parameter *φ*, with *ξ* = 0.9. Panels (a,b) correspond to 1*/α* = 6 months (*α* = 2, year^−1^), and panels (c,d) correspond to 1*/α* = 2 years (*α* = 0.5, year^−1^). As in Fig. 3, panels (a,c) show solution branches for the total infected population *I*, while panels (b,d) show corresponding extrema of ln(*I*). Labels indicate Hopf (*H*), pitchfork (*P* ), torus (*TR*), tangent/fold (*T* ), flip/period-doubling (*F* ), and boundary point (*BP* ) bifurcations, when present. The remaining parameters are fixed as in Table 2.

For the longer TCI duration, *α* = 0.5, changes in *φ* produce a wider region of complex dynamics. In particular, small and intermediate values of *φ* may generate irregular or multi-periodic oscillations before the system stabilizes at higher values. This indicates that the duration of TCI modulates the dynamical impact of homologous reinfection transmissibility. In epidemiological terms, even a relatively low contribution of homologous reinfections to onward transmission can interact with population immune structure and alter epidemic dynamics, whereas sufficiently high values of *φ* tend to stabilize endemic persistence under the baseline parameterization. Together, the *ξ* and *φ* bifurcation diagrams show that homologous susceptibility and homologous infectiousness affect dengue dynamics through different mechanisms, but that both effects are strongly conditioned by the duration of temporary cross-immunity.

The detailed boundary-branch structure associated with partly positive invariant subsystems is discussed in Appendix C. In particular, the analysis of the SIRSIR_1_ and SIRSIR_2_ subsystems clarifies how boundary points arise and how stability can be exchanged between single-strain invariant subsystems and the full coexistence dynamics. This technical discussion is separated from the main text to keep the Results focused on the epidemiological interpretation of the bifurcation diagrams.

#### 4.1.2 Two-parameter bifurcation diagrams

To examine how the duration of TCI interacts with homologous reinfection, we computed two-parameter bifurcation diagrams for the parameter pairs (*α, ξ*) and (*α, φ*), shown in Fig. 5(a) and Fig. 5(b), respectively. These diagrams summarize how changes in the TCI waning rate and homologous reinfection parameters structure the system’s qualitative dynamics. They therefore complement the one-parameter diagrams by assessing how the effects of homologous susceptibility and homologous infectiousness depend on the duration of cross-immunity.

**Figure 5:**
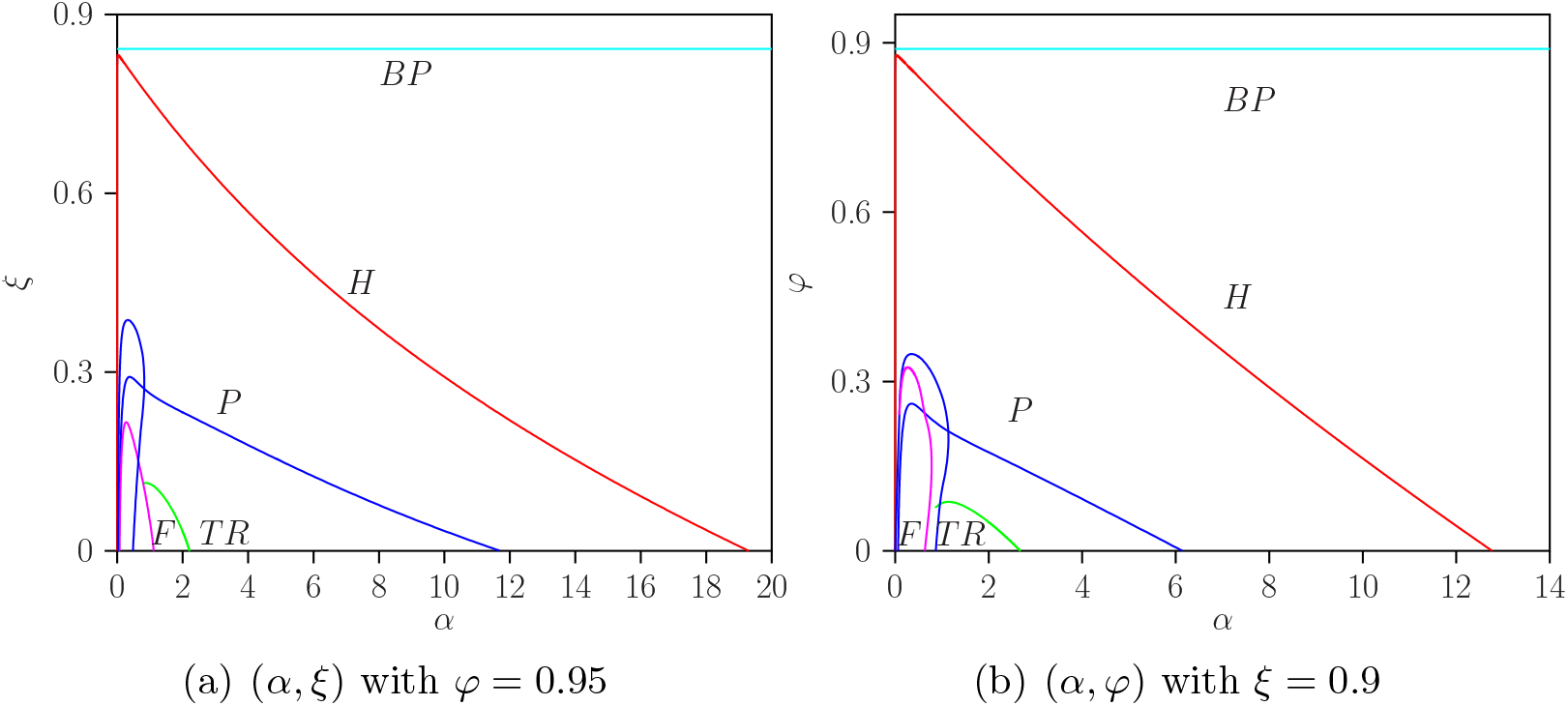
Two-parameter bifurcation diagrams for (a) (*α, ξ*) with *φ* = 0.95 and (b) (*α, φ*) with *ξ* = 0.9. Bifurcation labels follow the notation introduced in Fig. 3. The remaining parameters are fixed as in Table 2.

Both diagrams show that the qualitative dynamics are defined by the same bifurcation structures identified in the one-parameter diagrams. Above the *BP* curve and before the Hopf curve, the system admits either a stable endemic equilibrium or a partly positive equilibrium in which one strain pathway is absent. Between the Hopf and torus curves, stable and unstable limit cycles shape the oscillatory dynamics, with pitchfork bifurcations separating distinct periodic branches.

Although several flip bifurcations are present, they do not form a classical Feigenbaum cascade leading to chaos. Instead, the torus bifurcation curve lies near the boundary of parameter regions where irregular dynamics consistent with chaos can emerge, consistent with previous analyses of two-infection dengue models [3, 6]. In this sense, the two-parameter diagrams show that TCI duration modulates the dynamical impact of homologous reinfection: for some combinations of *α* and either *ξ* or *φ*, the system transitions from stable endemic persistence to periodic, quasi-periodic, or irregular dynamics consistent with chaos.

The diagrams for (*α, ξ*) and (*α, φ*) display broadly similar bifurcation structures, indicating that both susceptibility to homologous reinfection and transmissibility during homologous reinfection can reshape the long-term dynamics. However, the parameter regions associated with complex dynamics differ between the two diagrams. This distinction reflects the different epidemiological roles of *ξ* and *φ*: the former controls the probability that seropositive individuals acquire homologous reinfection, whereas the latter controls the infectious contribution of homologously reinfected individuals to onward transmission.

The presence of multiple branches and repeated stability changes suggests that, for some parameter combinations, different attractors may coexist and the long-term outcome may depend on the initial conditions. In these regions, stable endemic equilibria, periodic or quasi-periodic trajectories, and irregular dynamics may be reached from different initial distributions of immune histories or infected compartments. This possible multistability highlights the complexity of the extended dengue system and indicates that parameter values alone may not always uniquely determine the observed epidemic trajectory.

The special case *ξ* = *φ* = 0, corresponding to the original model without homologous reinfection, is discussed in Appendix B. In that limiting case, the dynamics are governed by the interaction between TCI and ADE-mediated differences in transmissibility during heterologous secondary infections, as in [3]. Comparing this limiting case with Fig. 5 shows how introducing homologous reinfection modifies, but does not replace, the dynamical mechanisms already present in the baseline two-infection model.

#### 4.1.3 Phase-space projections and time series for the non-seasonal model

The bifurcation diagrams are complemented by direct numerical simulations. Time series and phase-space projections provide representative trajectories for selected values of the homologous reinfection parameters and help illustrate the dynamical regimes identified by continuation analysis.

Figures 6 and 7 show representative trajectories for the intermediate TCI duration, 1*/α* = 6 months (*α* = 2 year^−1^). Figure 6 shows trajectories obtained by varying the homologous susceptibility parameter *ξ* while fixing *φ* = 0.95. For small values of *ξ*, the system displays irregular dynamics consistent with the chaotic regions identified in the bifurcation diagrams. As *ξ* increases, the trajectories become progressively more regular, passing through multi-periodic or double-cycle behavior before approaching a simpler periodic regime.

**Figure 6:**
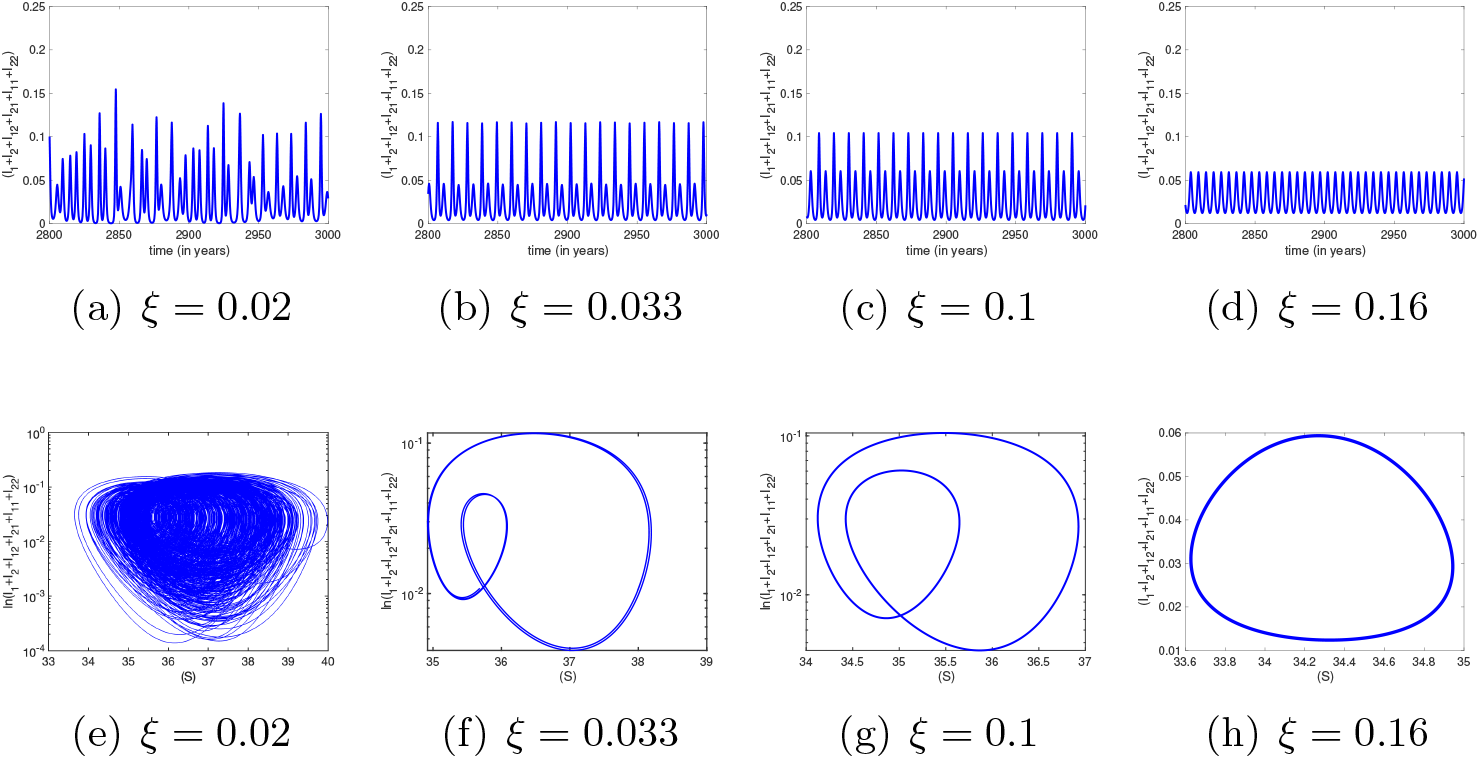
Representative time series (a–d) and corresponding phase-space projections (e–h) for the non-seasonal model with intermediate TCI duration, 1*/α* = 6 months (*α* = 2 year^−1^), obtained by varying the homologous susceptibility parameter *ξ* while fixing *φ* = 0.95.

**Figure 7:**
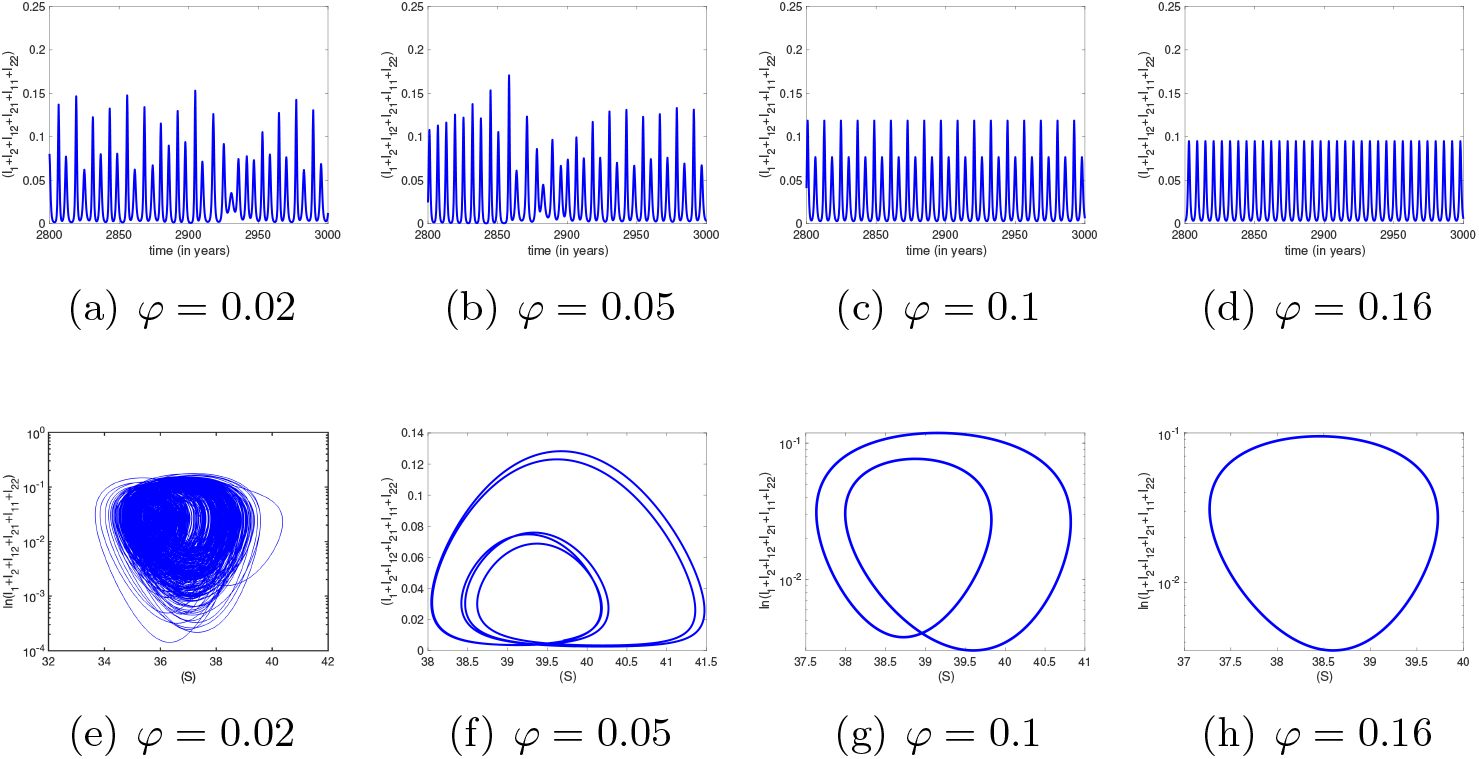
Representative time series (a–d) and corresponding phase-space projections (e–h) for the non-seasonal model with intermediate TCI duration, 1*/α* = 6 months (*α* = 2 year^−1^), obtained by varying the homologous transmissibility parameter *φ* while fixing *ξ* = 0.9.

Figure 7 shows the corresponding trajectories obtained by varying the homologous transmissibility parameter *φ* while fixing *ξ* = 0.9. These simulations illustrate a transition from irregular dynamics at low values of *φ* to more regular periodic behavior as *φ* increases. The sequence is consistent with the bifurcation diagrams in Fig. 4 and shows how increasing homologous transmissibility alters the system dynamics, with solutions transitioning from irregular regimes to periodic oscillations.

Figures 8 and 9 show the corresponding simulations for the longer TCI duration, 1*/α* = 2 years (*α* = 0.5 year^−1^). Compared with the intermediate TCI case, the longer period of cross-protection produces a wider range of complex oscillatory behavior. When *ξ* is varied with *φ* = 0.95, the system displays irregular dynamics consistent with chaos for low values of *ξ*, quasi-periodic behavior for intermediate values, and higher-period or more regular oscillations as *ξ* increases.

**Figure 8:**
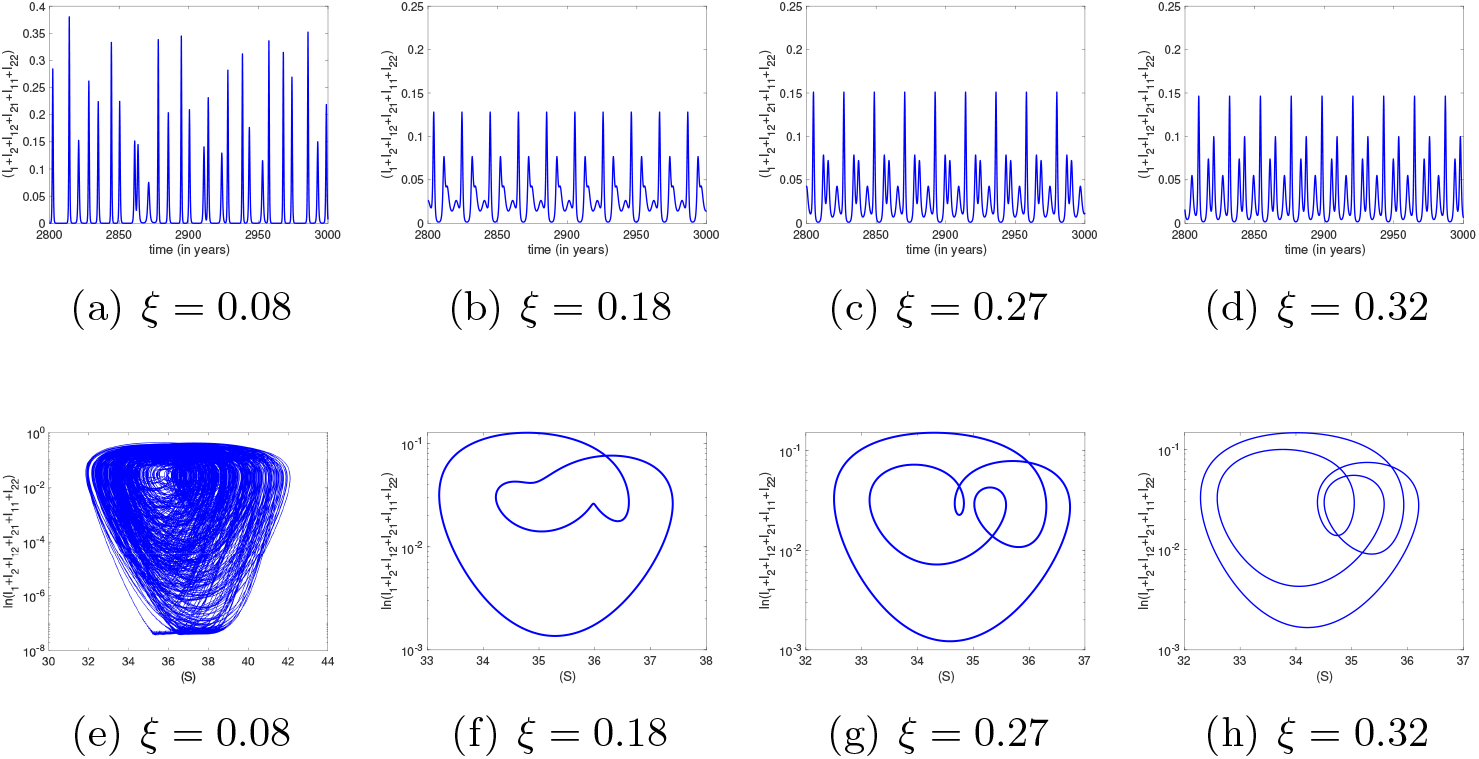
Representative time series (a–d) and corresponding phase-space projections (e–h) for the non-seasonal model with long TCI duration, 1*/α* = 2 years (*α* = 0.5 year^−1^), obtained by varying the homologous susceptibility parameter *ξ* while fixing *φ* = 0.95.

**Figure 9:**
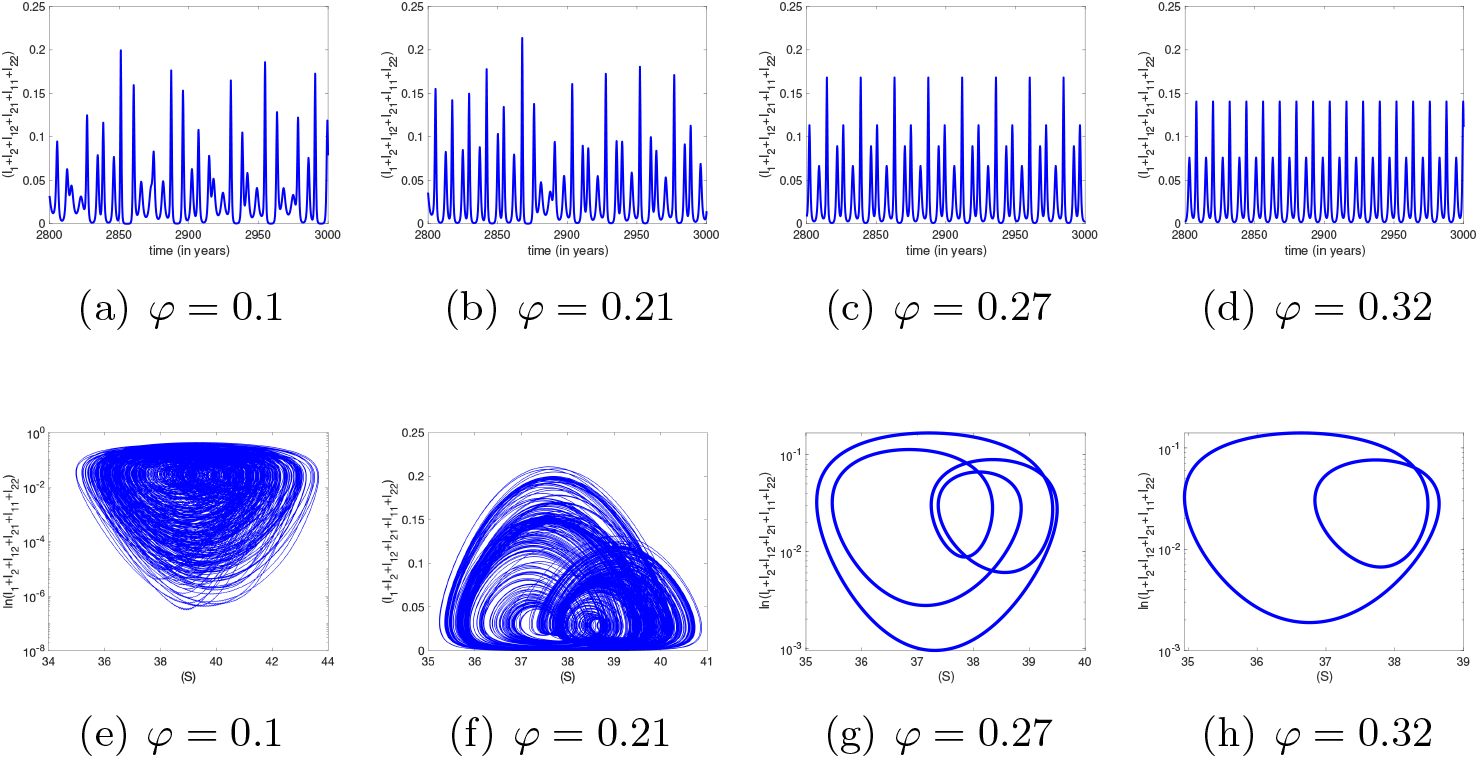
Representative time series (a–d) and corresponding phase-space projections (e–h) for the non-seasonal model with long TCI duration, 1*/α* = 2 years (*α* = 0.5 year^−1^), obtained by varying the homologous transmissibility parameter *φ* while fixing *ξ* = 0.9.

When *φ* is varied with *ξ* = 0.9, the system similarly passes through irregular, quasi-periodic, and multi-periodic regimes before approaching more regular oscillatory behavior, as shown in Fig. 9.

The time series and phase-space projections support the bifurcation results by showing that longer TCI can broaden the parameter region in which complex epidemic trajectories occur. In particular, the system may exhibit irregular outbreaks, quasi-periodic cycles, or multi-periodic oscillations depending on whether homologous reinfection is varied through susceptibility *ξ* or transmissibility *φ*. For the intermediate TCI duration, varying *ξ* or *φ* produces broadly similar transitions among irregular dynamics consistent with chaos, multi-periodic behavior, and periodic regimes. For the longer TCI duration, the two parameters have more distinct dynamical effects, indicating that TCI duration modulates whether homologous reinfection acts primarily through susceptibility to reinfection or through onward transmissibility. These simulations therefore complement the bifurcation analysis by illustrating how changes in *ξ, φ*, and *α* translate into observable epidemic time series and phase-space trajectories.

### 4.2 External forcing: seasonal variation in the transmission rate

To account for ecological and climatic drivers of mosquito abundance, such as temperature and rainfall, we introduce seasonal variation in the effective transmission rate. Since seasonal changes in mosquito abundance affect biting rates and, consequently, dengue transmission, we replace the constant transmission parameter *β* by

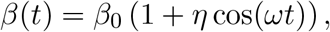

where *ω* = 2*π/T* and *T* = 1 year. The parameter *η* controls the amplitude of seasonal forcing. With this external forcing, the model becomes a non-autonomous ODE system.

Following previous work [3, 49], we consider a high seasonal forcing scenario with *η* = 0.35. We then analyze how the homologous susceptibility parameter *ξ* and the homologous transmissibility parameter *φ* shape the dynamics under seasonal forcing. This provides an additional sensitivity assessment of homologous reinfection parameters in the presence of an external annual transmission cycle.

Figures 10 and 11 show one-parameter bifurcation diagrams for the seasonally forced model with intermediate TCI duration, 1*/α* = 6 months (*α* = 2 year^−1^), and long TCI duration, 1*/α* = 2 years (*α* = 0.5 year^−1^), respectively. We consider *ξ* ∈ [0, 1] and *φ* ∈ [0, 1], using the total infected population

**Figure 10:**
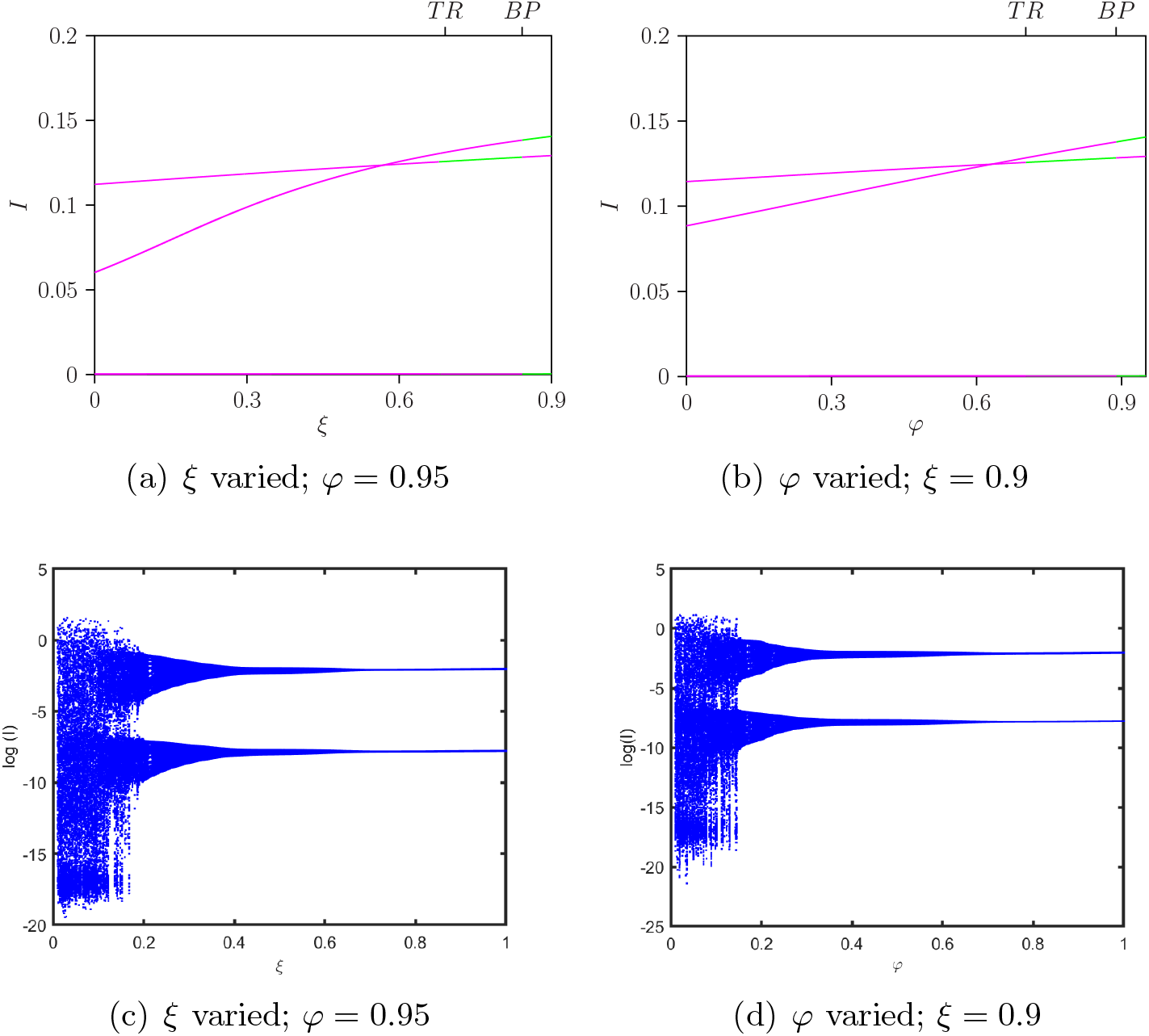
One-parameter bifurcation diagrams for the seasonally forced model with *η* = 0.35 and intermediate TCI duration, 1*/α* = 6 months (*α* = 2 year^−1^). Panels (a,c) vary the homologous susceptibility parameter *ξ* with *φ* = 0.95, whereas panels (b,d) vary the homologous transmissibility parameter *φ* with *ξ* = 0.9. Panels (a,b) show solution branches for the total infected population *I*, while panels (c,d) show the corresponding extrema of ln(*I*). The remaining parameters are fixed as in Table 2.

**Figure 11:**
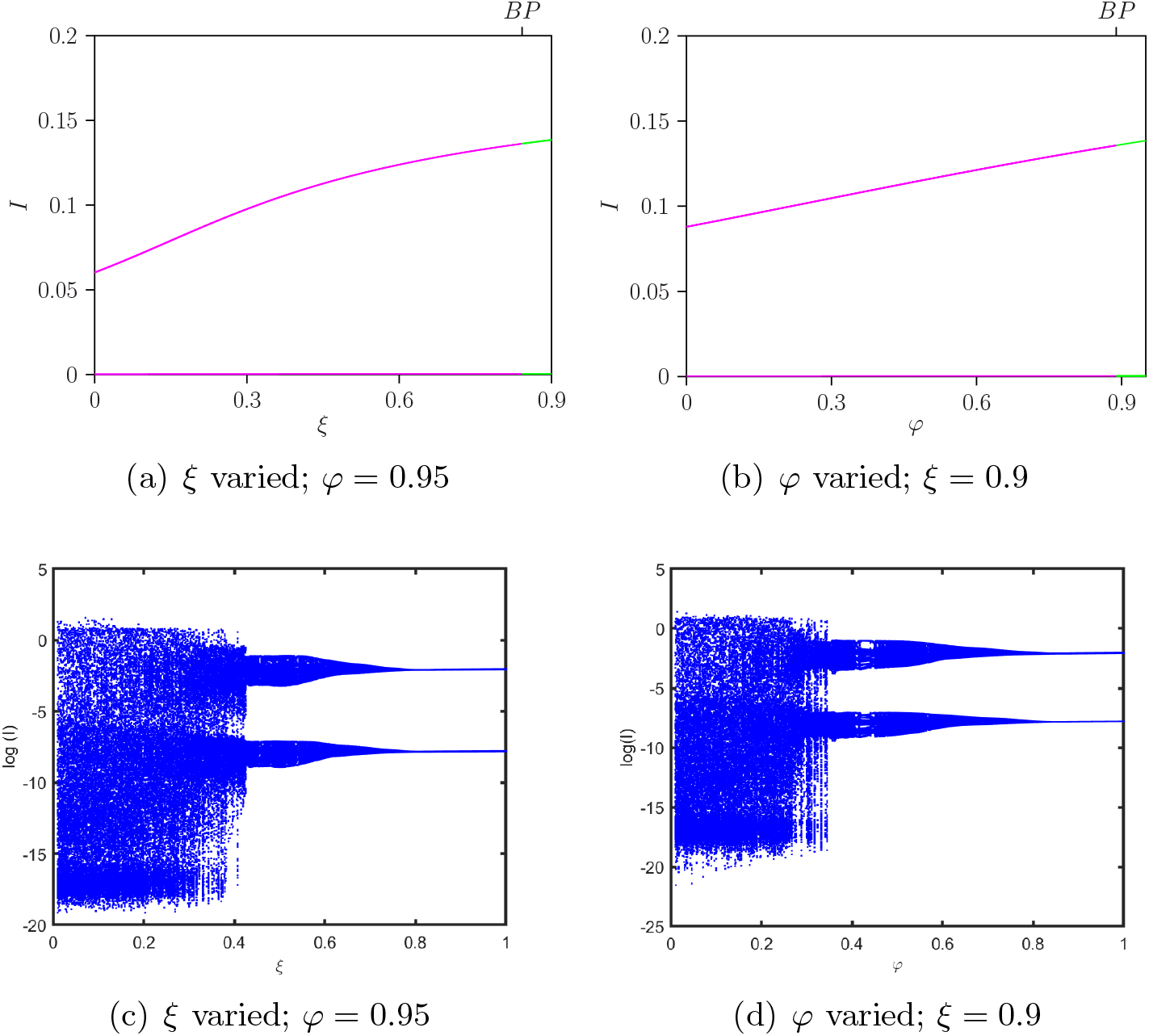
One-parameter bifurcation diagrams for the seasonally forced model with *η* = 0.35 and long TCI duration, 1*/α* = 2 years (*α* = 0.5 year^−1^). Panels (a,c) vary the homologous susceptibility parameter *ξ* with *φ* = 0.95, whereas panels (b,d) vary the homologous transmissibility parameter *φ* with *ξ* = 0.9. Panels (a,b) show solution branches for the total infected population *I*, while panels (c,d) show the corresponding extrema of ln(*I*). The remaining parameters are fixed as in Table 2.

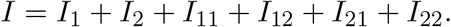

Compared with the non-seasonal case, seasonal forcing alters the bifurcation structure and reshapes the distribution of oscillatory regimes. For both TCI durations and for variations in either *ξ* or *φ*, the diagrams show that seasonal forcing shifts the location of bifurcation points and modifies the parameter regions associated with periodic and more complex oscillatory dynamics. The comparison between *α* = 2 and *α* = 0.5 shows that the effect of homologous reinfection under seasonal forcing remains strongly conditioned by TCI duration.

Because the forced system is high-dimensional and non-autonomous, different dynamical regimes may coexist for the same parameter values. In such cases, the long-term trajectory can depend on the initial distribution of susceptible, infected, and immune individuals. Thus, seasonal forcing may not only shift bifurcation curves, but may also increase sensitivity to initial conditions by allowing different periodic, quasi-periodic, or irregular attractors to be reached under the same epidemiological parameters.

#### 4.2.1 Two-parameter bifurcation diagrams with seasonal forcing

To assess how seasonal forcing modifies the interaction between TCI duration and homologous reinfection, we computed two-parameter bifurcation diagrams for the pairs (*α, ξ*) and (*α, φ*) under high seasonal forcing, *η* = 0.35 (Fig. 12). In contrast to the non-seasonal system, the dominant structures are boundary points (*BP* ) and torus bifurcations (*TR*).

**Figure 12:**
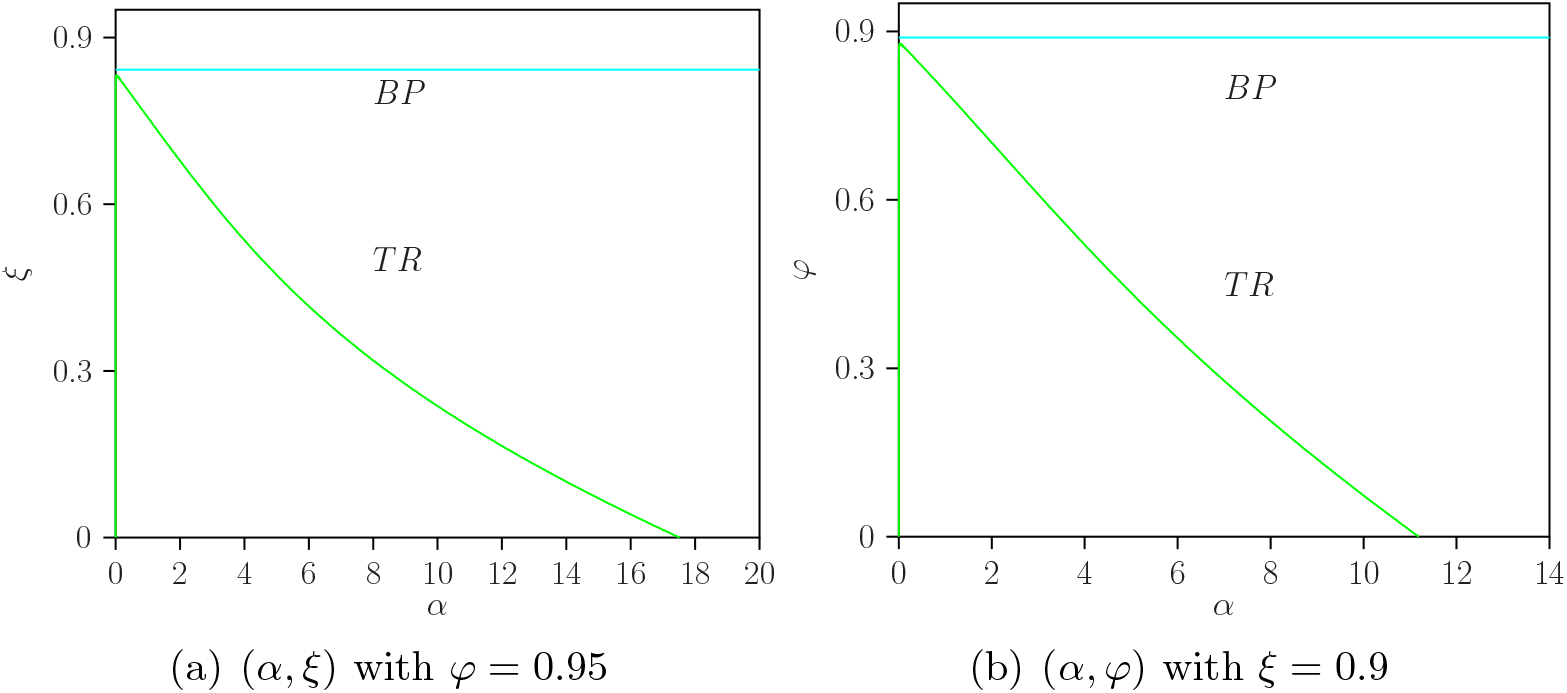
Two-parameter bifurcation diagrams for the seasonally forced model with *η* = 0.35: (a) (*α, ξ*) with *φ* = 0.95, and (b) (*α, φ*) with *ξ* = 0.9. The main bifurcation curves are boundary points (*BP* ) and torus bifurcations (*TR*). The remaining parameters are fixed as in Table 2.

The *BP* curve separates parameter regions in which partly positive dynamics occur from regions in which full two-strain coexistence persists. Because the system is periodically forced, endemic states are represented by periodic solutions rather than stationary equilibria. Between the *BP* and *TR* curves, the dynamics are shaped by stable and unstable periodic solutions. Beyond the *TR* curve, the system may exhibit quasi-periodic behavior or irregular dynamics consistent with chaos. Thus, under seasonal forcing, torus bifurcations become a central route through which homologous reinfection and TCI duration jointly generate complex epidemic trajectories.

The presence of multiple branches and repeated stability changes suggests that, in some parameter regions, different long-term trajectories may coexist for the same parameter values. In such cases, the observed epidemic trajectory may depend on the initial distribution of susceptible, infected, and immune individuals. This possible multistability highlights the complexity of the seasonally forced system and indicates that parameter values alone may not always uniquely determine the observed dynamical regime.

Representative time series and phase-space projections for the seasonally forced model are provided in Appendix A. These simulations illustrate the same qualitative transitions shown by the bifurcation analysis while keeping the main text focused on the dominant bifurcation structure.

## 5 Discussion

In this study, we extended a two-infection multi-strain dengue model to investigate how homologous reinfection may affect long-term epidemic dynamics. Classical multi-strain dengue models usually assume that infection with a given serotype confers complete lifelong protection against reinfection by the same serotype. Motivated by recent empirical and modeling evidence suggesting that homologous reinfections can occur, we relaxed this assumption and introduced two parameters describing homologous reinfection: the relative susceptibility to homologous reinfection, *ξ*, and the relative infectiousness of homologously reinfected individuals, *φ*. This allowed us to examine how homologous reinfection interacts with temporary cross-immunity (TCI), ADE-mediated differences in transmission, and seasonal forcing.

The results show that homologous reinfection can substantially reshape the qualitative dynamics of dengue transmission, even when susceptibility to homologous reinfection is reduced relative to primary infection. In the non-seasonal model, varying either *ξ* or *φ* produced transitions among stable endemic equilibria, periodic oscillations, quasi-periodic behavior, and irregular dynamics consistent with chaos. These transitions were strongly modulated by the duration of TCI. For an intermediate TCI duration, corresponding to an average cross-protection period of six months, the system displayed windows of irregular dynamics when homologous susceptibility was low, followed by periodic regimes and eventually stable endemic persistence as *ξ* increased. For a longer TCI duration, corresponding to an average cross-protection period of two years, the parameter regions associated with complex oscillatory behavior became broader. This suggests that longer-lasting cross-protection can amplify the dynamical effects of even weak homologous reinfection pathways.

The distinction between *ξ* and *φ* is epidemiologically important. The parameter *ξ* controls the probability that a seropositive individual becomes reinfected with the same serotype, whereas *φ* controls the contribution of homologously reinfected individuals to onward transmission. Although both parameters affect the same reinfection pathway, they act at different stages of transmission. Our bifurcation analysis shows that changes in susceptibility and changes in infectiousness can generate broadly similar qualitative regimes, but the location and extent of the periodic, quasi-periodic, and irregular regions differ. Thus, the epidemiological impact of homologous reinfection depends not only on whether such reinfections occur, but also on how infectious they are and how they interact with the immune structure of the host population.

A central implication of the analysis is that homologous reinfection should be modeled as a rare but dynamically relevant pathway. Small values of *ξ*, corresponding to strong but incomplete serotype-specific protection, are sufficient to reproduce complex qualitative dynamics observed in endemic dengue settings, including recurrent outbreaks, multi-periodic oscillations, and irregular dynamics consistent with chaos. In contrast, larger values of *ξ* tend to stabilize transmission toward endemic persistence and are therefore less consistent with the observed irregularity of dengue incidence. Thus, the most epidemiologically relevant regime is not one in which homologous reinfection is frequent, but one in which low-probability homologous reinfection interacts with TCI and ADE-mediated transmission differences to sustain complex epidemic patterns.

A key finding is that the introduction of homologous reinfection does not create an entirely new dynamical mechanism independent of the previously known TCI–ADE interaction. Rather, it modifies and reshapes the bifurcation structure already present in two-infection dengue models. In the limiting case *ξ* = *φ* = 0, the model reduces to the previously studied framework in which complex dynamics arise from the interaction between TCI and differences in transmission between primary and heterologous secondary infections. When homologous reinfection is introduced, the baseline structure is preserved but modified: bifurcation thresholds shift, oscillatory windows expand or contract, and the routes to irregular dynamics can change. This supports the interpretation that homologous reinfection acts as an additional immune-transmission feedback rather than as a replacement for the classical mechanisms underlying complex dengue dynamics.

The analysis of endemic equilibria also revealed the mathematical possibility of backward bifurcation and bistability when *ξ >* 1. This result should be interpreted with caution. In the present biological interpretation, *ξ* represents relative susceptibility to homologous reinfection after prior infection with the same serotype. Therefore, biologically meaningful values satisfy 0 ≤ *ξ* ≤ 1, and values above one imply enhanced susceptibility to homologous reinfection, which is not consistent with the assumed partial protection conferred by serotype-specific immunity. Consequently, the backward bifurcation obtained for *ξ >* 1 should not be interpreted as evidence that homologous reinfection generates bistability under realistic dengue parameter values. Instead, it identifies a mathematical mechanism by which sufficiently strong reinfection feedback could create multiple endemic equilibria if homologous reinfection were artificially amplified. Within the biologically relevant range considered here, the main epidemiological role of homologous reinfection lies in its effect on oscillatory, quasi-periodic, and irregular epidemic dynamics, rather than in generating biologically realistic backward bifurcation.

Seasonal forcing further modified the dynamical landscape. By allowing the transmission rate to vary periodically, we introduced a simple representation of ecological and climatic effects on mosquito abundance and biting rates. Under high seasonal forcing, the model became non-autonomous and the bifurcation structure changed substantially. In particular, torus bifurcations became a central mechanism for complex trajectories. While the non-seasonal model exhibited complex dynamics through combinations of Hopf, pitchfork, flip, and torus bifurcations, the seasonally forced model showed that external periodic forcing can shift the system toward quasiperiodic and irregular regimes. This indicates that seasonality can amplify the effects of homologous reinfection, especially when combined with longer periods of TCI.

The results also suggest possible multistability in some regions of parameter space. Because the system is high-dimensional and contains several immune-history compartments, different attractors may coexist for the same parameter values. In such cases, the observed long-term trajectory may depend on the initial distribution of susceptible, infected, temporarily immune, and seropositive individuals. This dependence on initial conditions is epidemiologically relevant because endemic populations with similar transmission parameters may differ substantially in their immune-history structure, serotype exposure, and recent epidemic history. Therefore, the model should be interpreted not only in terms of parameter-driven transitions, but also as a framework in which initial immune conditions can influence the epidemic regime reached by the system.

From an epidemiological perspective, these results highlight the importance of immune history in dengue transmission. Even if homologous reinfections are rare at the individual level, their population-level effects may be non-negligible when they interact with cross-immunity, ADE-mediated transmission differences, and seasonal variation. In highly exposed populations, repeated immune stimulation, waning cross-protection, and partial susceptibility to reinfection may alter outbreak timing, recurrence, and amplitude. These effects are particularly relevant for interpreting irregular dengue incidence patterns in endemic regions and for understanding how serotype-specific immunity shapes long-term transmission.

## Limitations

The model has several limitations. First, we considered only two explicit strains, following previous work showing that the interaction between TCI and ADE can drive rich dynamics even in reduced strain systems. This provides a parsimonious framework for bifurcation analysis, but it does not capture all possible interactions among the four dengue serotypes. Future extensions could include all four serotypes, asymmetric serotype-specific transmission, and differences in immune protection or enhancement across serotype pairs.

Second, vector dynamics were represented implicitly through effective transmission parameters. This simplification is useful for identifying the nonlinear effects of immune history, homologous reinfection, and seasonality, but it does not capture explicit mosquito population dynamics, vector control, climate-dependent vector life-history traits, or spatial variation in vector abundance. Including explicit vector compartments would allow the model to distinguish between human immune mechanisms and ecological drivers of transmission, such as temperature-dependent biting rates, mosquito mortality, extrinsic incubation, and intervention-driven changes in vector density.

Third, the model assumes homogeneous mixing. This assumption may affect the quantitative interpretation of the results, since dengue transmission is often shaped by age, household structure, local mobility, spatial clustering of vectors, and heterogeneous exposure to mosquito bites. Age structure could modify the distribution of immune histories and the timing of primary and secondary infections. Spatial heterogeneity could generate asynchronous local outbreaks, travelling waves, or source–sink dynamics that are not represented in the present well-mixed framework. Behavioural heterogeneity, including changes in mobility, care-seeking, or vector-avoidance behaviour during outbreaks, may also alter the effective contribution of different infection classes to onward transmission.

Fourth, homologous reinfection was represented phenomenologically through the parameters *ξ* and *φ*. More detailed clinical, virological, and epidemiological data on the frequency, viral load, duration of viraemia, symptom severity, and mobility patterns of homologously reinfected individuals would be needed to estimate these parameters in specific settings. Until such data become available, the values of *ξ* and *φ* should be interpreted as exploratory sensitivity parameters rather than directly measured biological constants.

Fifth, although we performed systematic one- and two-parameter bifurcation analyses, we did not perform a full global sensitivity analysis across all epidemiological parameters. Such an analysis would be useful for ranking the relative importance of ADE strength, TCI duration, seasonal forcing amplitude, transmission intensity, demographic turnover, and homologous reinfection parameters for specific quantitative outcomes. Future work could combine bifurcation analysis with Latin-hypercube sampling, PRCC, Sobol sensitivity indices, or uncertainty quantification methods to assess the robustness of the dynamical regimes identified here.

Finally, the identification of irregular dynamics was based on bifurcation structure, time series, and phase-space projections. Future work could complement these analyses with quantitative diagnostics such as Lyapunov exponents, entropy-based measures, recurrence analysis, or data-assimilation approaches applied to long-term dengue incidence time series.

## 6 Conclusions

Relaxing the assumption of complete homologous protection enriches the dynamical behavior of multi-strain dengue models. In this framework, small values of *ξ*, corresponding to strong but incomplete serotype-specific protection, are sufficient to reproduce complex qualitative dynamics observed in endemic dengue settings, whereas larger values of *ξ* tend to stabilize transmission and are less consistent with irregular dengue incidence. These findings emphasize that homologous reinfection is best interpreted as a rare but dynamically influential pathway and that dengue models should account for immune history and reinfection pathways when investigating long-term transmission dynamics. The results also indicate possible multistability in the extended model, with different long-term trajectories attainable for the same parameter values depending on the initial distribution of susceptible, infected, and immune individuals. This dependence on initial conditions is epidemiologically relevant because endemic populations can differ substantially in immune-history structure, even when transmission parameters are similar.

Future work should connect this theoretical framework with serological, virological, and cohort data to better quantify homologous reinfection parameters and assess their relevance across epidemiological settings. Further extensions should also incorporate explicit vector dynamics, age and spatial structure, behavioural heterogeneity, and global sensitivity analysis to evaluate the robustness of the dynamical regimes identified here.

## Data Availability

Not applicable

## Declarations

### Ethics approval and consent to participate

Not applicable.

### Consent for publication

Not applicable.

### Data availability

No new empirical data were generated or analyzed in this study. All parameter values used in the numerical simulations are provided in the manuscript.

### Materials availability

Not applicable.

### Code availability

The code used for numerical simulations and bifurcation analysis is available from the corresponding author upon reasonable request.

### Author contributions

A.K.S., V.S., N.S., B.W.K., and M.A. contributed to the conception and design of the study. A.K.S., and B.W.K. performed the mathematical analysis, numerical simulations, and bifurcation analysis. V.S., N.S., and M.A. contributed to model interpretation and validation of the dynamical-systems results. A.K.S., V.S., and M.A. drafted the manuscript. All authors contributed to manuscript revision and approved the final version.

## A Additional results for the seasonally forced model

For completeness, we provide additional bifurcation diagrams and representative trajectories for the seasonally forced model. These supplementary results support the main-text conclusion that seasonal forcing reshapes the bifurcation structure and can make torus bifurcations a central route to complex epidemic dynamics.

### A.1 Two-parameter bifurcation diagrams with seasonal forcing

To assess how seasonal forcing modifies the interaction between TCI duration and homologous reinfection, we computed two-parameter bifurcation diagrams for the pairs (*α, ξ*) and (*α, φ*) under high seasonal forcing, *η* = 0.35 (Fig. 13). In contrast to the non-seasonal system, the dominant bifurcation structures are boundary points (*BP* ) and torus bifurcations (*TR*).

**Figure 13:**
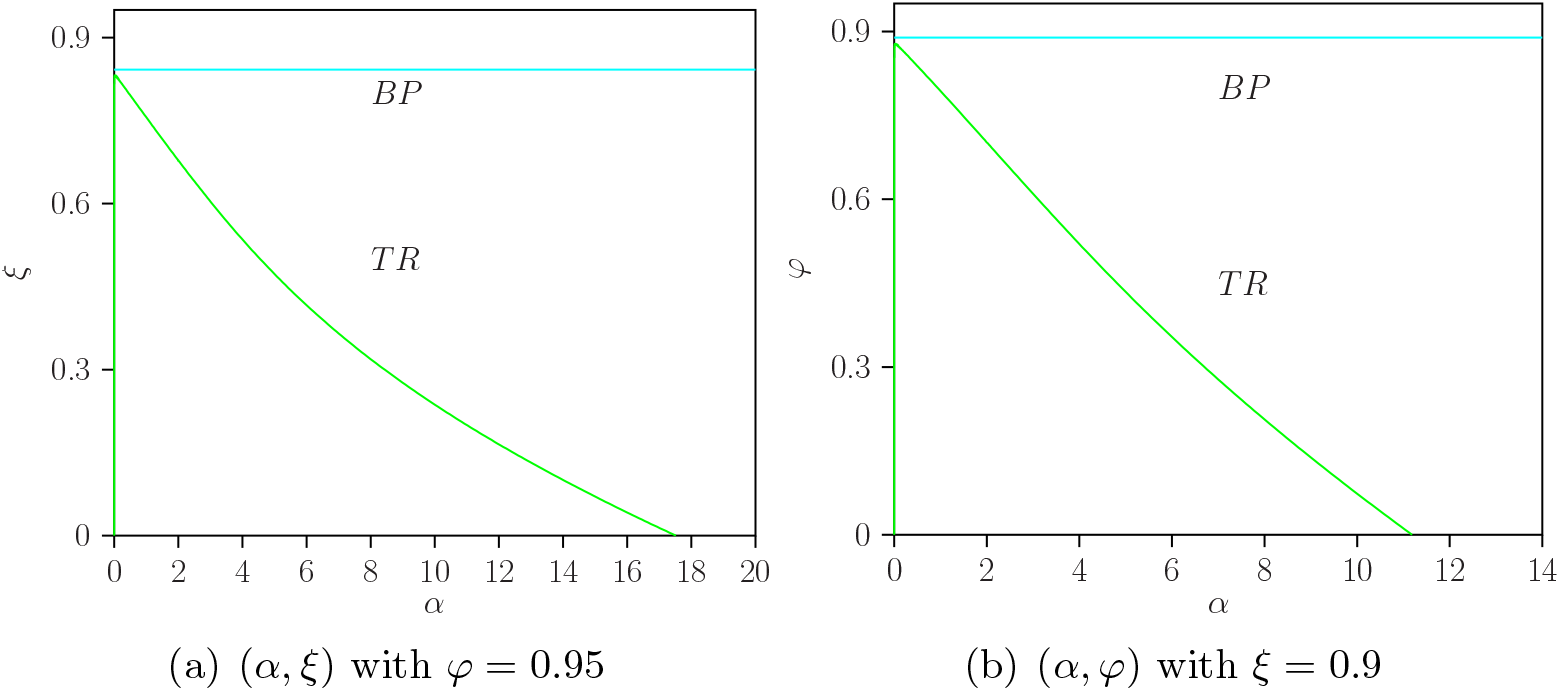
Two-parameter bifurcation diagrams for the seasonally forced model with *η* = 0.35: (a) (*α, ξ*) with *φ* = 0.95, and (b) (*α, φ*) with *ξ* = 0.9. The main bifurcation curves are boundary points (*BP* ) and torus bifurcations (*TR*). The remaining parameters are fixed as in Table 2.

The presence of multiple branches and repeated stability changes also suggests possible multistability in some regions of parameter space. In such cases, different long-term trajectories may occur for the same parameter values depending on the initial distribution of susceptible, infected, and immune individuals. This possible dependence on initial conditions is consistent with the high-dimensional and non-autonomous structure of the seasonally forced system.

### A.2 Phase-space projections and time series for the seasonal model

We next provide representative trajectories of the seasonally forced model. Figures 14 and 15 correspond to the intermediate TCI duration, 1*/α* = 6 months (*α* = 2, year^−1^). Figures 16 and 17 show the corresponding simulations for the longer TCI duration, 1*/α* = 2 years (*α* = 0.5, year^−1^).

**Figure 14:**
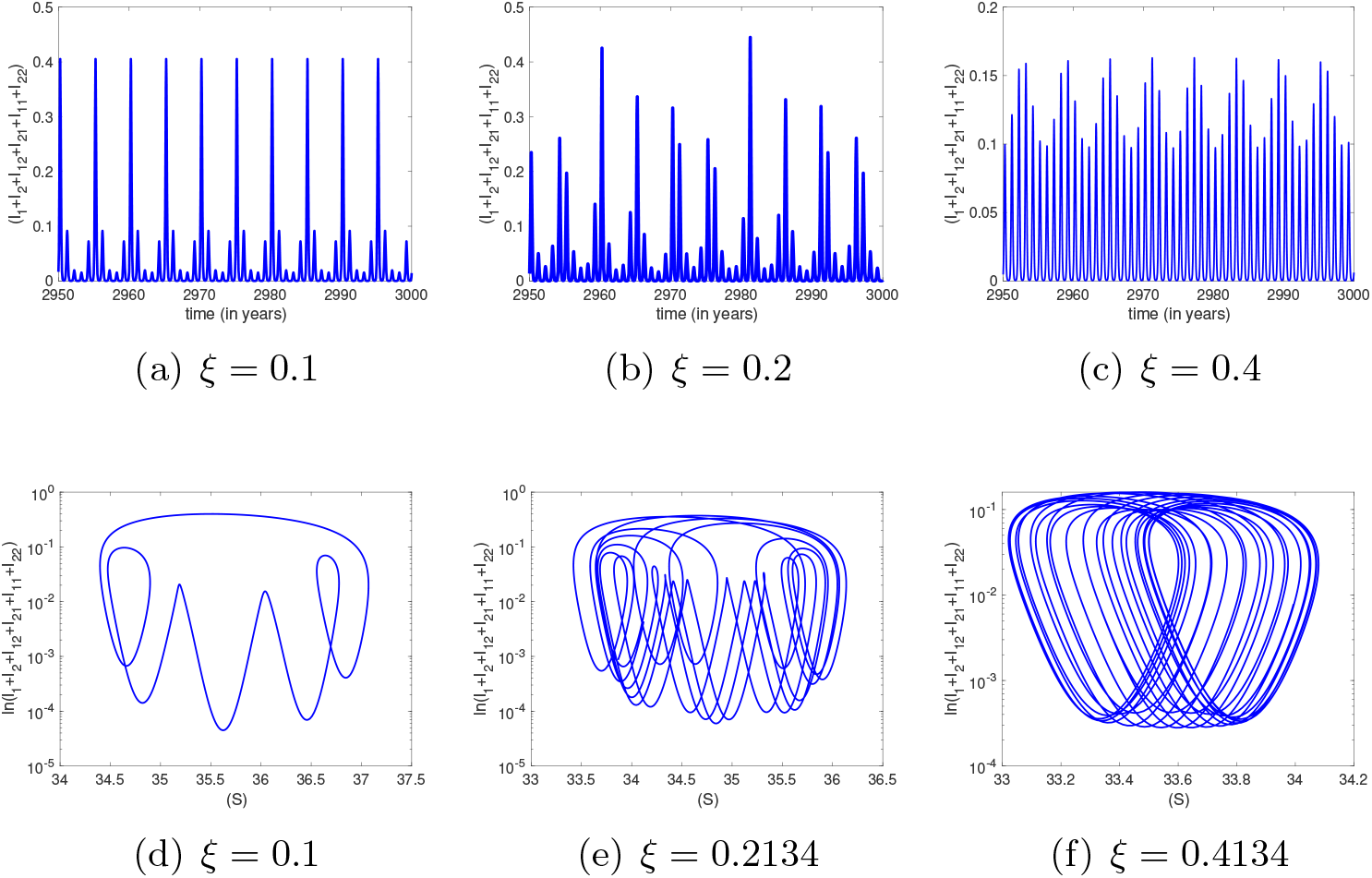
Representative time series (a–c) and corresponding phase-space projections (d–f) for the seasonally forced model with intermediate TCI duration, 1*/α* = 6 months (*α* = 2, year^−1^), obtained by varying *ξ* while fixing *φ* = 0.95 and *η* = 0.35.

**Figure 15:**
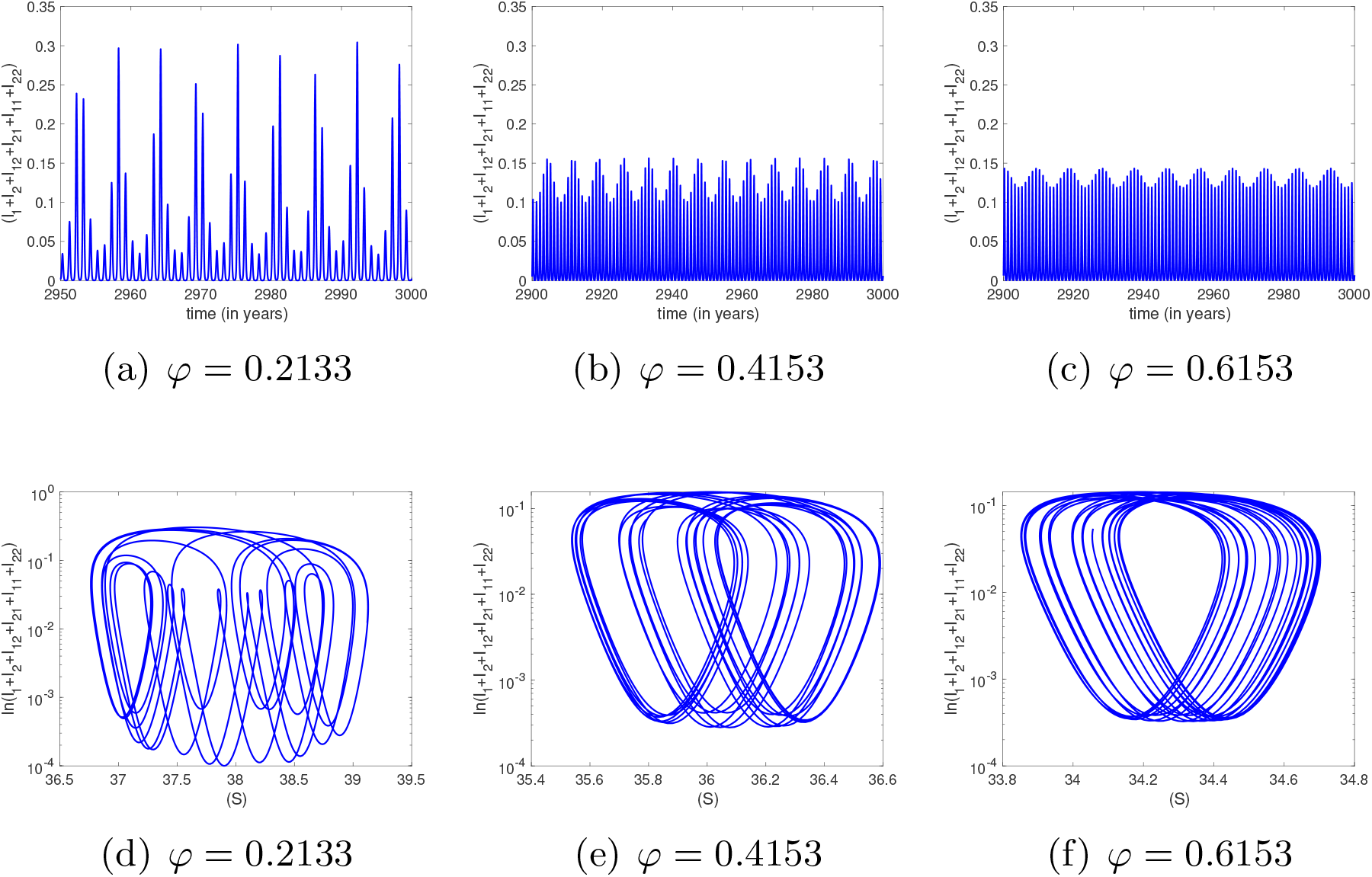
Representative time series (a–c) and corresponding phase-space projections (d–f) for the seasonally forced model with intermediate TCI duration, 1*/α* = 6 months (*α* = 2, year^−1^), obtained by varying *φ* while fixing *ξ* = 0.9 and *η* = 0.35.

**Figure 16:**
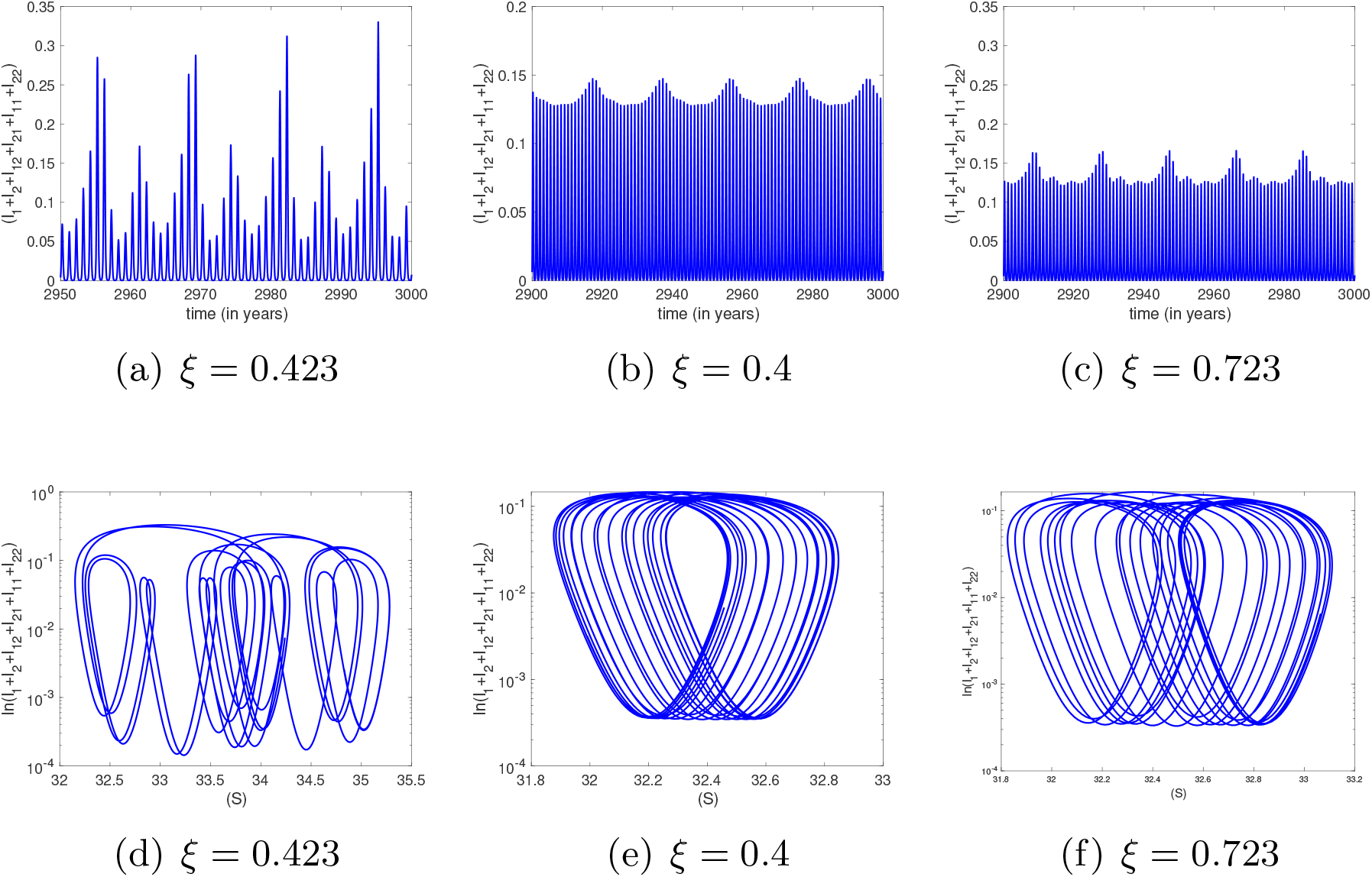
Representative time series (a–c) and corresponding phase-space projections (d–f) for the seasonally forced model with long TCI duration, 1*/α* = 2 years (*α* = 0.5, year^−1^), obtained by varying *ξ* while fixing *φ* = 0.95 and *η* = 0.35.

**Figure 17:**
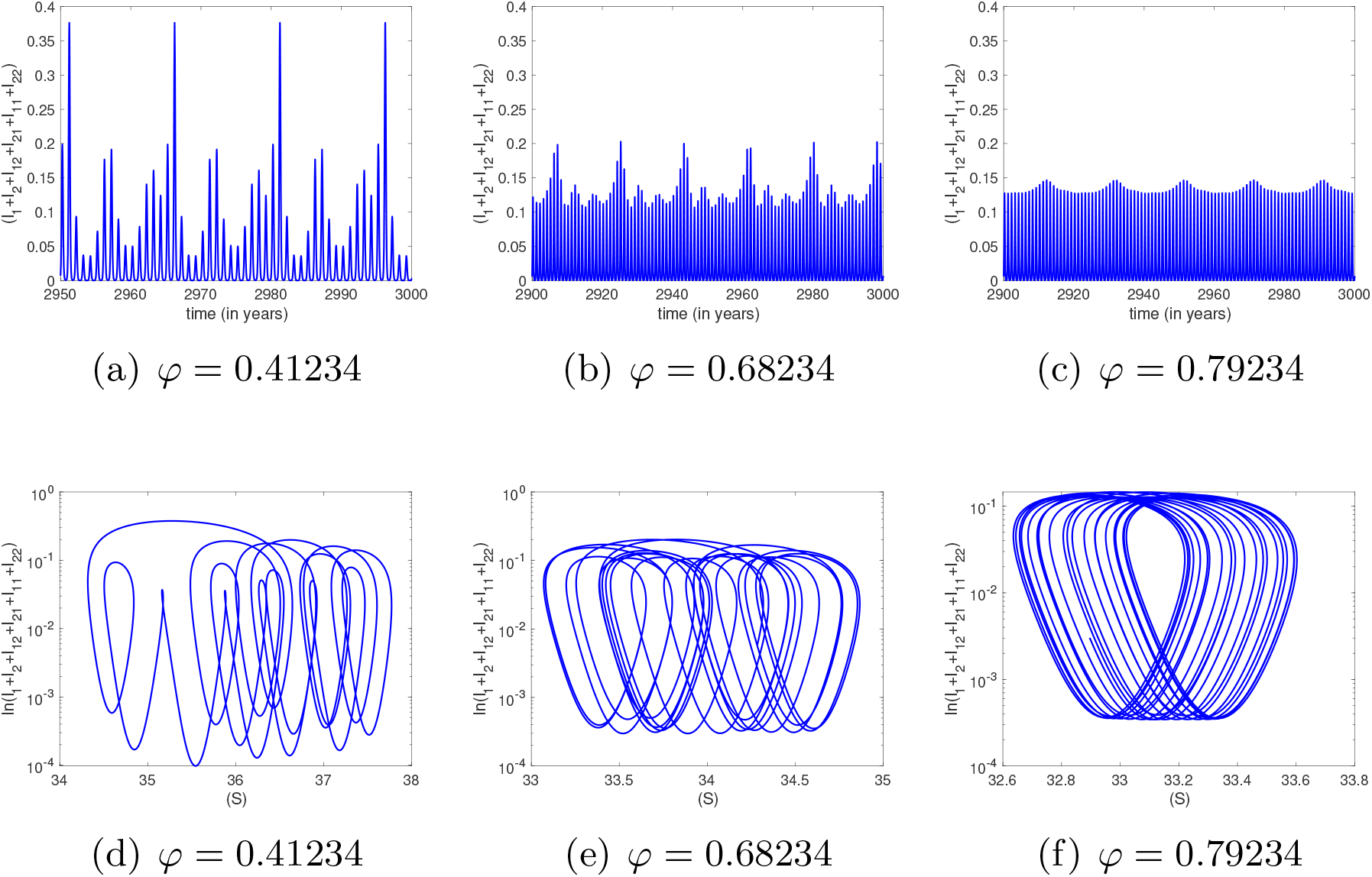
Representative time series (a–c) and corresponding phase-space projections (d–f) for the seasonally forced model with long TCI duration, 1*/α* = 2 years (*α* = 0.5, year^−1^), obtained by varying *φ* while fixing *ξ* = 0.9 and *η* = 0.35.

For the intermediate TCI duration, varying *ξ* with *φ* = 0.95 shows a progression from irregular dynamics consistent with chaos toward torus-organized oscillations as *ξ* increases. A similar transition is observed when varying *φ* with *ξ* = 0.9, although the parameter values at which these regimes occur differ. This indicates that seasonal forcing changes the route to complex dynamics compared with the non-seasonal model.

For the longer TCI duration, seasonal forcing again produces complex trajectories, but the parameter ranges associated with irregular, quasi-periodic, and multi-periodic behavior are shifted. This is consistent with the two-parameter diagrams, which show that TCI duration interacts with homologous reinfection parameters in organizing the transition between regular and irregular epidemic trajectories.

Together, these supplementary simulations show that seasonal forcing modifies the route to complex dynamics. In the non-seasonal system, complex behavior arises through combinations of Hopf, pitchfork, flip, and torus bifurcations. Under high seasonal forcing, torus bifurcations become a dominant mechanism, and irregular dynamics consistent with chaos emerge beyond these torus curves. Thus, seasonal fluctuations in transmission can amplify or reshape the dynamical effects of homologous reinfection.

## B Limiting case without homologous reinfection

When *ξ* = *φ* = 0, homologous reinfection is removed from the model. In this limiting case, the system reduces to the previously studied two-infection dengue framework with TCI and ADE-mediated differences between primary and heterologous secondary infections [1, 3]. This comparison is useful because it separates the dynamical mechanisms already present in the baseline model from those introduced by homologous reinfection.

The corresponding two-parameter bifurcation diagram in the (*ϕ, α*) plane is shown in Fig. 18, while one-parameter bifurcation diagrams obtained by varying *ϕ* are shown in Fig. 19. These diagrams recover the rich dynamics described in previous studies, including coexistence of stable limit cycles, tangent/fold bifurcations, flip bifurcations, torus bifurcations, and irregular dynamics consistent with chaos.

**Figure 18:**
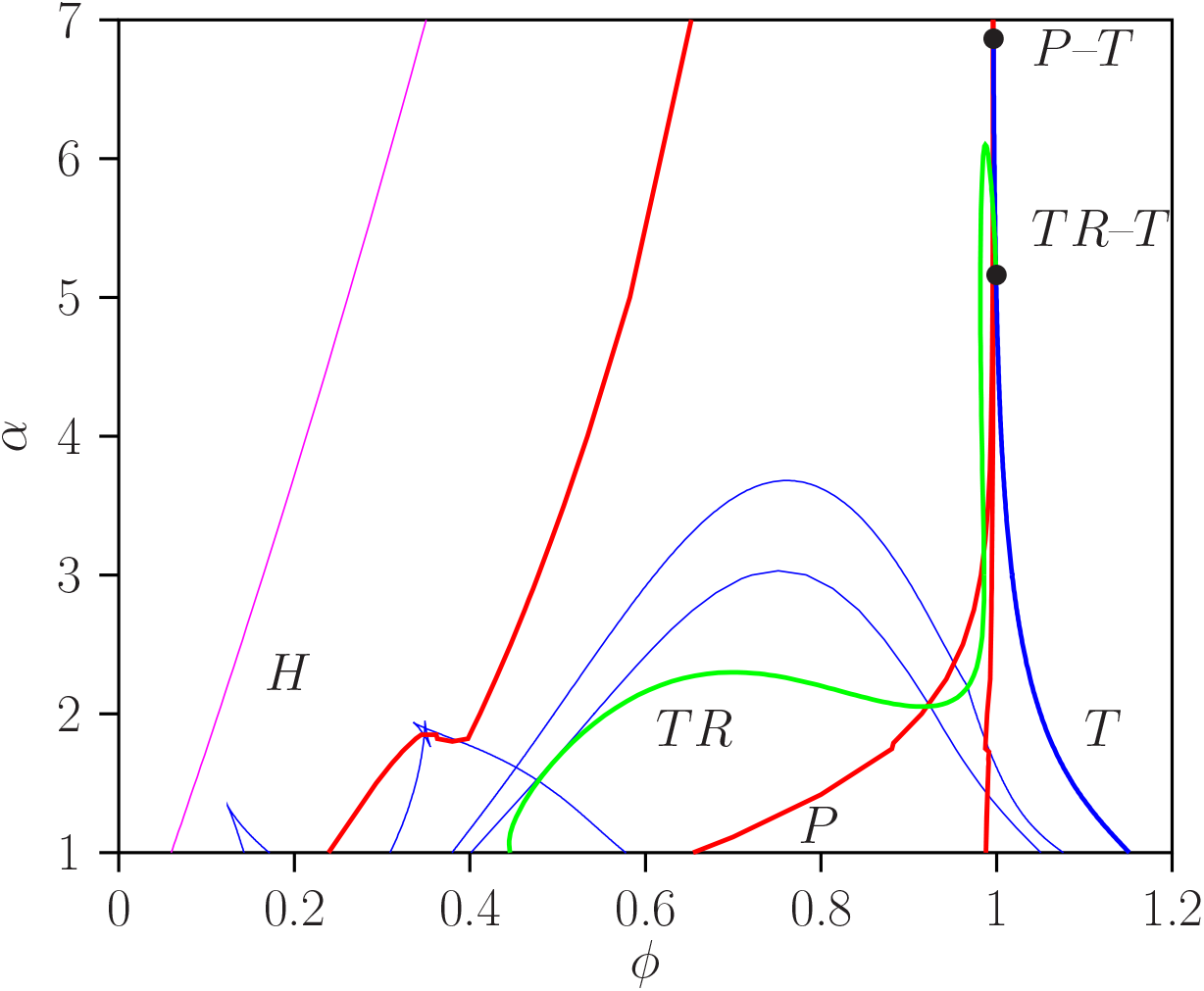
Two-parameter bifurcation diagram for the limiting case *ξ* = *φ* = 0, corresponding to the model without homologous reinfection. The diagram is shown in the (*ϕ, α*) parameter plane. Bifurcation curves include Hopf bifurcations (*H*), tangent/fold bifurcations (*T* ), flip or period-doubling bifurcations (*F* ), and torus bifurcations (*TR*), when present.

**Figure 19:**
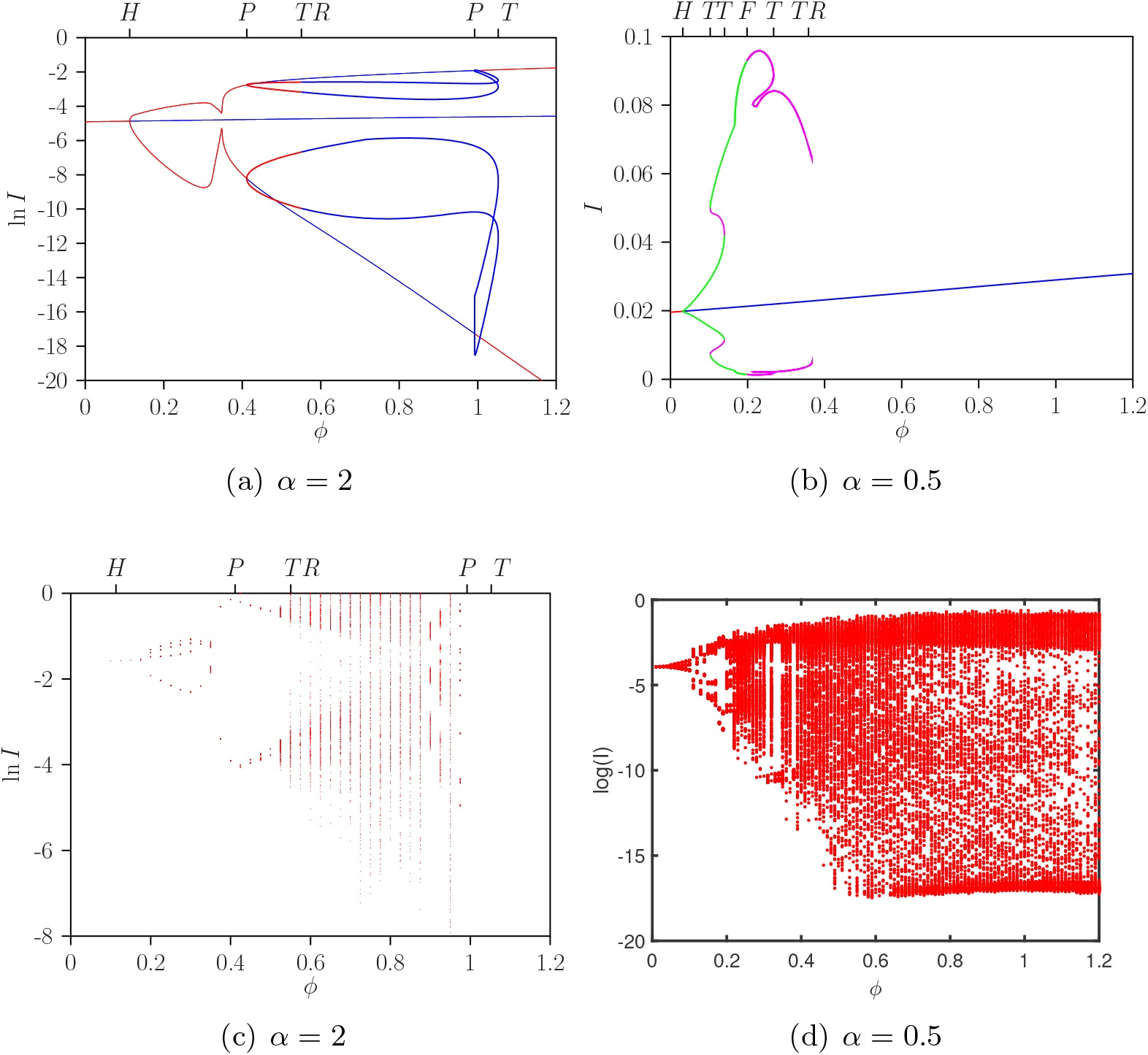
One-parameter bifurcation diagrams for the limiting case *ξ* = *φ* = 0, obtained by varying the heterologous secondary infectiousness parameter *ϕ*. Panels (a,c) correspond to 1*/α* = 6 months (*α* = 2, year^−1^), and panels (b,d) correspond to 1*/α* = 2 years (*α* = 0.5, year^−1^). Panels (a,b) show solution branches for the total infected population, while panels (c,d) show corresponding extrema. These diagrams are consistent with the bifurcation structure previously reported for the two-infection dengue model without homologous reinfection [1, 3].

**Figure 20:**
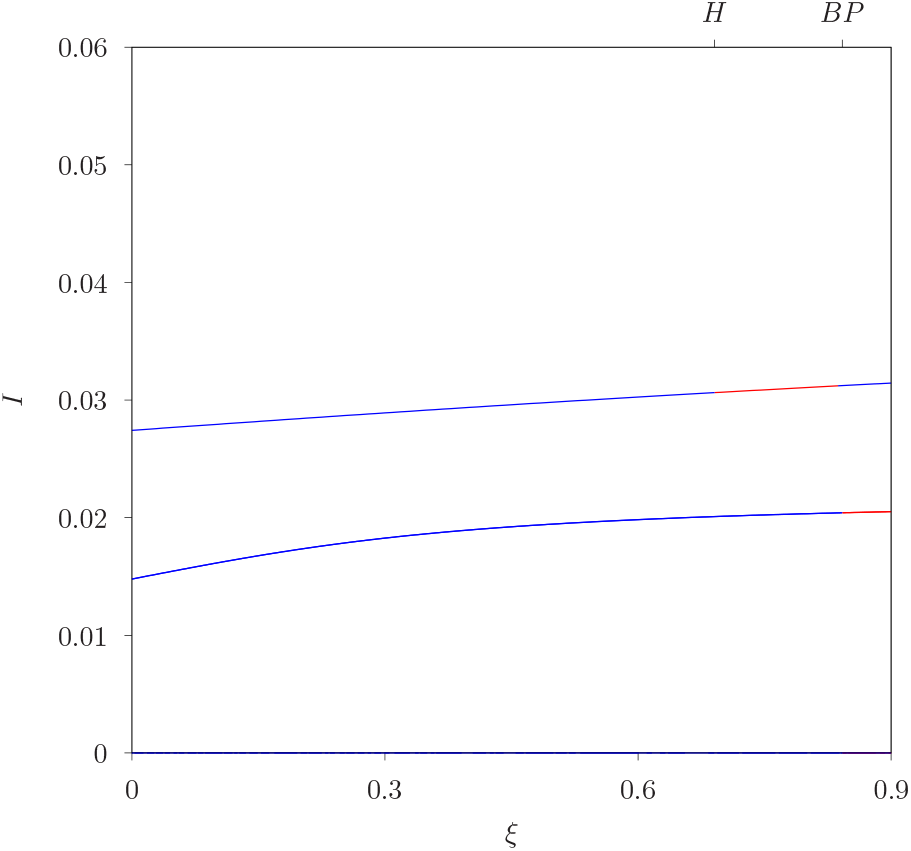
Detail of the bifurcation structure for the state variable *I* with *φ* = 0.95, showing solution branches associated with the partly positive subsystems SIRSIR_1_ and SIRSIR_2_.

## C Boundary subsystem structure and Filippovtype analogy

To better understand the boundary bifurcation structure observed in the one-parameter diagrams, we examined two partly positive invariant subsystems corresponding to dominance of one strain pathway. These subsystems arise when one strain pathway is absent, while the other strain and its homologous reinfection pathway remain active.

The first subsystem is

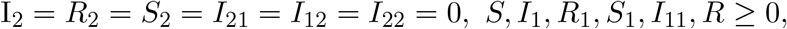

and the second subsystem is

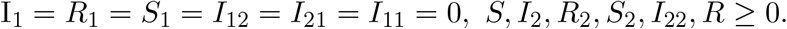

These subsystems correspond to trajectories in which only one primary strain pathway and its homologous reinfection pathway remain active.

For lower values of the homologous reinfection parameter, the partly positive subsystems SIRSIR_1_ and SIRSIR_2_ interact with the full system near the boundary point denoted by *BP* . At this point, the stability of the full endemic equilibrium and the partly positive branches changes. Although this behavior is reminiscent of boundary bifurcations studied in piecewise-smooth or Filippov systems [19, 11], the present model is smooth throughout the state space. Therefore, the analogy is qualitative: the boundary point reflects an exchange of stability between invariant subsystems and the full coexistence dynamics, rather than a discontinuity in the vector field.

## D Additional bifurcation diagrams for variation in *ϕ*

For completeness, we also report additional one-parameter bifurcation diagrams obtained by varying the heterologous secondary infectiousness parameter *ϕ* under different external forcing assumptions. These diagrams compare the baseline non-seasonal model, the model with imported infections, and the model with both imported infections and low seasonal forcing. Here, *ρ* denotes the rate of imported infections used in the supplementary comparison. This appendix is included only to document additional robustness checks from the broader modelling framework. These simulations are not used to support the main conclusions of the present manuscript, which focus on homologous reinfection through *ξ* and *φ*.

Figure 22 shows that the inclusion of imported infections can stabilize parts of the dynamics by reducing or removing irregular windows observed in the non-seasonal system. When low seasonal forcing is added together with importation, irregular dynamics may reappear over a wider range of *ϕ*. These results are included only as a supplementary comparison, since the main text focuses on the effects of homologous reinfection through *ξ* and *φ*.

**Figure 21:**
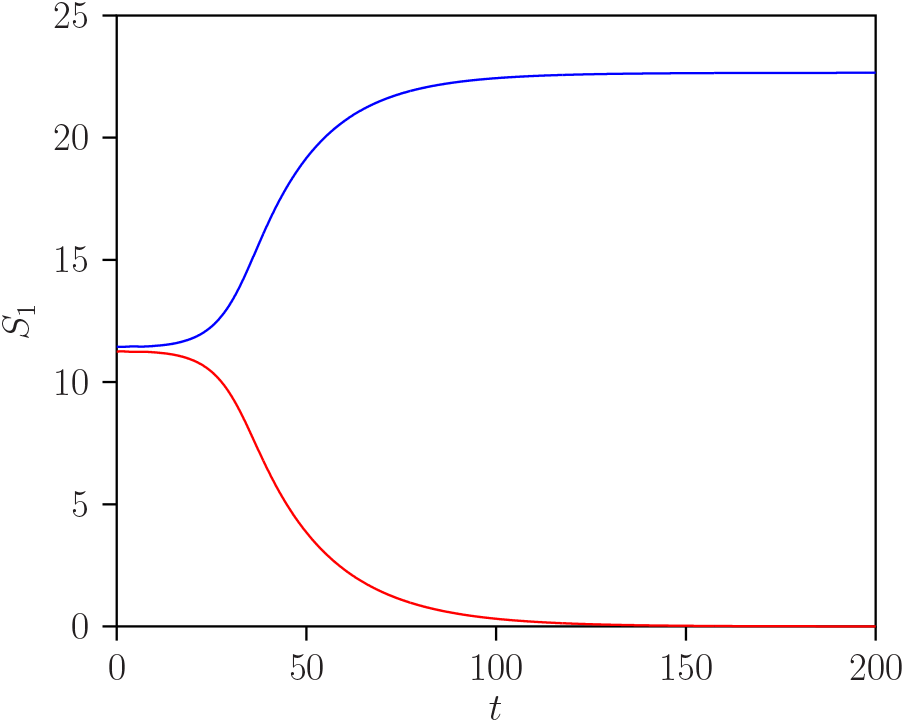
Time evolution of *S*_1_ for initial conditions above and below the *S*_1_ equilibrium value of the full system (2.1), with *φ* = 0.95. The black curve corresponds to the SIRSIR_1_ subsystem and the red curve to the SIRSIR_2_ subsystem.

**Figure 22:**
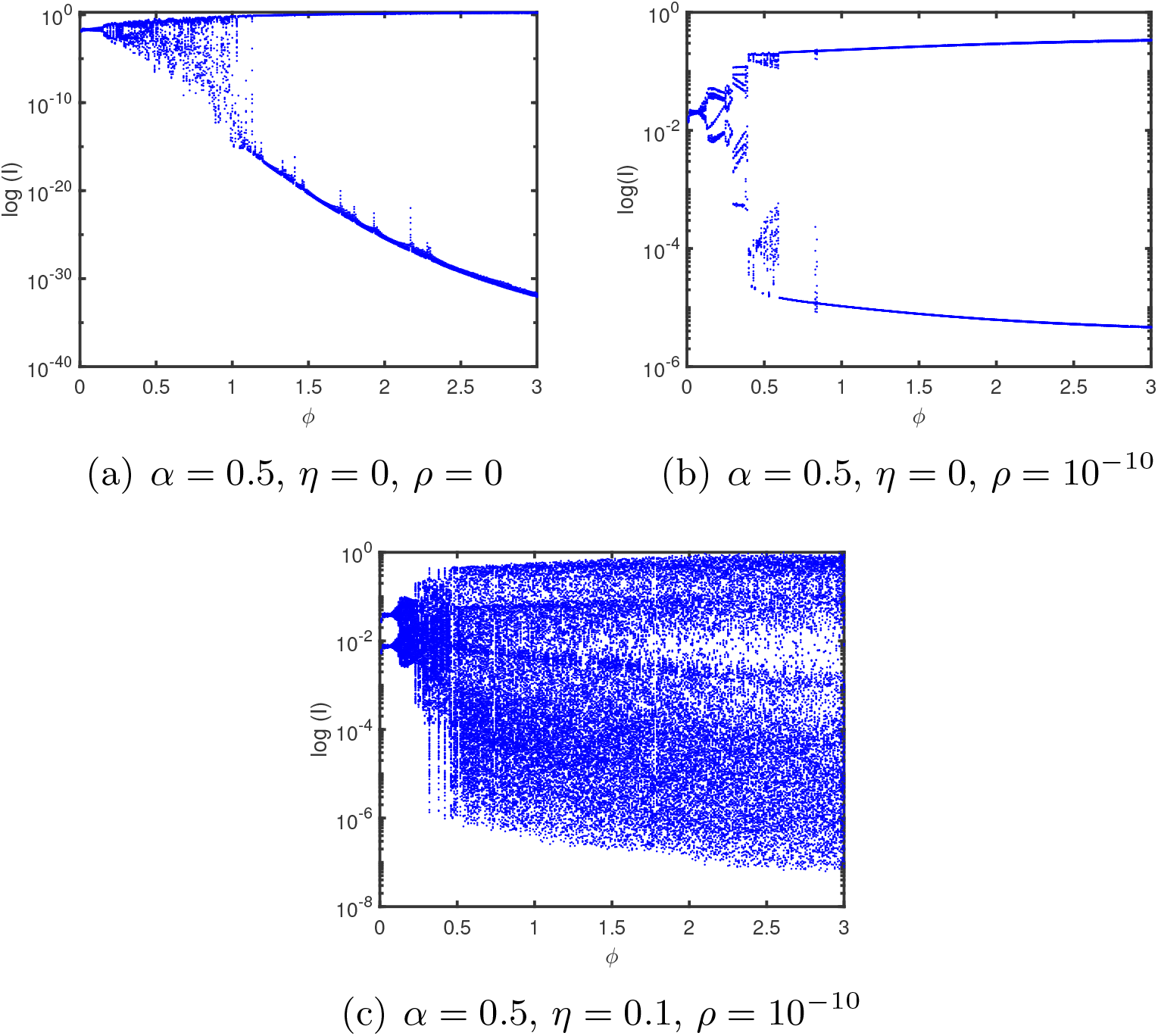
Additional one-parameter bifurcation diagrams obtained by varying the heterologous secondary infectiousness parameter *ϕ* under different external forcing assumptions. Panel (a) shows the non-seasonal model without importation, panel (b) includes importation, and panel (c) includes both importation and low seasonal forcing. Other fixed parameters include *β* = 2*γ, γ* = 52, year^−1^, *µ* = 1*/*65, year^−1^, and *φ* = 0.95.

## References

[1] Aguiar M., Kooi B., Stollenwerk N. (2008) Epidemiology of Dengue Fever: A Model with Temporary Cross-Immunity and Possible Secondary Infection Shows Bifurcations and Chaotic Behaviour in Wide Parameter Regions. Mathematical Modelling of Natural Phenomena. 3(4):48–70. 10.1051/mmnp:2008070

[2] Aguiar M., Anam V., Blyuss K.B., et al. (2002) Mathematical models for dengue fever epidemiology: A 10-year systematic review. Phys Life Rev. 40:65–92. 10.1016/j.plrev.2022.02.001

[3] Aguiar M., Ballesteros S., Kooi B. W., Stollenwerk N. (2011) The role of seasonality and import in a minimalistic multi-strain dengue model capturing differences between primary and secondary infections: Complex dynamics and its implications for data analysis. Journal of Theoretical Biology. 289:181—196.

[4] Aguiar M., Kooi B. W., Rocha F., et al. (2013) How much complexity is needed to describe the fluctuations observed in dengue hemorrhagic fever incidence data? Ecological Complexity. 16:31—40.

[5] Aguiar M., Stollenwerk N., Kooi B. W. (2009) Torus bifurcations, isolas and chaotic attractors in a simple dengue fever model with ADE and temporary cross immunity. International Journal of Computer Mathematics. 86(10–11):1867—1877.

[6] Aguiar M., Steindorf V., Srivastav A.K., et al. (2024) Bifurcation analysis of a two infection SIR-SIR epidemic model with temporary immunity and disease enhancement. Nonlinear Dyn. 112: 13621—13639 10.1007/s11071-024-09710-9

[7] Anam V., Sebayang A. A., Fahlena H., Knopoff D., Stollenwerk N., Soewono E., Aguiar M. (2022) Modeling Dengue Immune Responses Mediated by Antibodies: Insights on the Biological Parameters to Describe Dengue Infections. Computational and Mathematical Methods. 2022:1–11. https://www.hindawi.com/journals/cmm/2022/8283239/

[8] Anam V., Guerrero B.V., Srivastav A.K., Stollenwerk N., Aguiar M. (2024) Within-host models unravelling the dynamics of dengue reinfections. Infectious Disease Modelling. 9(2):458–473. https://www.sciencedirect.com/science/article/pii/S2468042724000150

[9] Aogo R.A., et al. (2023) Effects of boosting and waning in highly exposed populations on dengue epidemic dynamics. Sci. Transl. Med. 1734. https://doi.org/110.1126/scitranslmed.adi1734

[10] Bhatt P., Sabeena S.P., Varma M., Arunkumar G. (2021) Current Understanding of the Pathogenesis of Dengue Virus Infection. Curr Microbiol. 78(1):17–32. 10.1007/s00284-020-02284-w

[11] Biák M., Hanus T., Janovská D. (2013) Some applications of Filippov’s dynamical systems. Journal of Computational and Applied Mathematics. 254:132–143.

[12] Corbett K.S., Katzelnick L., Tissera H., Amerasinghe A., de Silva A.D., de Silva A.M. (2015) Preexisting neutralizing antibody responses distinguish clinically inapparent and apparent dengue virus infections in a Sri Lankan pediatric cohort. J Infect Dis. 211(4):590–599. 10.1093/infdis/jiu481

[13] Dhooge A., Govaerts W., Kuznetsov Yu. A. (2003) MATCONT: A MATLAB package for numerical bifurcation analysis of ODEs. ACM Trans Math Software. 29(2):141—64.

[14] Doedel E.J., Fairgrieve T.F., Sandstede B., Champneys A.R., Kuznetsov Y.A., Wang X. (2007) AUTO-07P: Continuation and bifurcation software for ordinary differential equations. Technical report, 2007.

[15] European Centre for Disease Prevention and Control (ECDC). Local transmission of dengue virus in mainland EU/EEA. https://www.ecdc.europa.eu/en/all-topics-z/dengue/surveillance-and-disease-data/autochthonous-transmission-dengue-virus-eueea

[16] European Centre for Disease Prevention and Control (ECDC). Dengue worldwide overview. https://www.ecdc.europa.eu/en/dengue-monthly

[17] Ferguson N.M., et al. (1999) The effect of antibody-dependent enhancement on the transmission dynamics and persistence of multiple-strain pathogens Proc. Natl. Acad. Sci. U.S.A.

[18] Ferguson N.M., et al. (1999) Transmission dynamics and epidemiology of dengue: insights from age-stratified sero-prevalence surveys Phil. Trans. Roy. Soc. Lond. B.

[19] Filippov A.F. (1964) Differential equations with discontinuous righthand side. In American Mathematical Society Translations. Series 2, AMS, Ann Arbor, pp. 199–231.

[20] Forshey B.M., Stoddard S.T., Morrison A.C. (2016) Dengue Viruses and Lifelong Immunity: Reevaluating the Conventional Wisdom. The Journal of Infectious Diseases. 214(7):979—981. 10.1093/infdis/jiw102

[21] Gubler D.J. (1998) Dengue and Dengue Hemorrhagic Fever. Clin Microbiol Rev. 11: 480—496.

[22] Guerrero B.V., Steindorf V., Blasco-Aguado R., Mateus L., Cevidanes A., et al. (2025) Assessing the spatio-temporal risk of Aedes-borne arboviral diseases in non-endemic regions: The case of Northern Spain. PLOS Neglected Tropical Diseases 19(7): e0013325. 10.1371/journal.pntd.0013325

[23] Guzman M.G., Gubler D.J., Izquierdo A., et al. (2016) Dengue infection. Nat Rev Dis Primers. 2 (1):16055. http://www.nature.com/articles/nrdp201655

[24] Guzmán M.G., Kouri G. (2002) Dengue: an update. Lancet Infect Dis. 2(1):33–42. 10.1016/S1473-3099(01)00171-2

[25] Halstead S.B., Udomsakdi S., Simasthien P., Singharaj P., Sukhavachana P., Nisalak A. (1970) Observations related to pathogenesis of dengue hemorrhagic fever. I. Experience with classification of dengue viruses. Yale J Biol Med. 42(5):261–275.

[26] Halstead S.B. (2002) Dengue hemorrhagic fever: two infections and antibody dependent enhancement, a brief history and personal memoir Rev Cuba Med Trop. 54:171–179. http://scielo.sld.cu/scielo.php?

[27] Halstead S.B. (2003) Neutralization and antibody-dependent enhancement of dengue viruses Advances in virus research. vol. 60. Academic Press pp: 421–467. https://www.sciencedirect.com/science/article/abs/pii/S0065352703600114

[28] Kathryn B. Anderson K.B., Gibbons R.V., Cummings D.A.T, et al. (2014) A Shorter Time Interval Between First and Second Dengue Infections Is Associated With Protection From Clinical Illness in a School-based Cohort in Thailand. The Journal of Infectious Diseases. 209(3):360–368. 10.1093/infdis/jit436

[29] Kooi B.W., Aguiar M., Stollenwerk N. (2014) Analysis of an asymmetric two-strain dengue model. Math Biosci. 10.1016/j.mbs.2013.12.009.

[30] MATLAB. Version 7.10.0 (R2010a). Natick, Massachusetts: The Math-Works Inc. 2010.

[31] Mier-y-Teran-Romero L., Schwartz I. B., Cummings D. A. T. (2013) Breaking the symmetry: Immune enhancement increases persistence of dengue viruses in the presence of asymmetric transmission rates. Journal of Theoretical Biology. 332:203–210. 10.1016/j.jtbi.2013.04.036

[32] Montoya M., Gresh L., Mercado J.C., Williams K.L., Vargas M.J., et al. (2013) Symptomatic Versus Inapparent Outcome in Repeat Dengue Virus Infections Is Influenced by the Time Interval between Infections and Study Year. PLOS Neglected Tropical Diseases 7(8): e2357. 10.1371/journal.pntd.0002357

[33] Naveca, F. G., Santiago, G. A., Maito, R. M., et al. (2023) Reemergence of Dengue Virus Serotype 3, Brazil, 2023. Emerging infectious diseases, 29(7), 1482–1484. 10.3201/eid2907.230595

[34] Patel B., Longo P., Miley M.J., Montoya M., Harris E., de Silva A.M. (2017) Dissecting the human serum antibody response to secondary dengue virus infections. PLoS Negl Trop Dis. 11(5):e0005554. 10.1371/journal.pntd.0005554

[35] Pisaneschi G., Manfredi P., Landi A., Stollenwerk N., Aguiar M. (2026). When Few Mosquitoes Are Enough: Dengue outbreaks in non-endemic areas, One Health. 22. 2026. 10.1016/j.onehlt.2025.101308

[36] Roussel M.R. (2019) Bifurcation analysis with AUTO. In: Nonlinear dynamics. Morgan & Claypool Publishers. pp. 2053–571, 5–1 to 5–13.

[37] Sabin A.B. (1952) Research on dengue during World War II. Am J Trop Med Hyg. 1:30–50. 10.4269/ajtmh.1952.1.30

[38] Sacchetto L., Bernardi V., Brancini M.L. (2025) Early insights of dengue virus serotype 3 (DENV-3) re-emergence in São Paulo, Brazil. Journal of Clinical Virology. 176, 105763. 10.1016/j.jcv.2025.105763

[39] Sangkawibha N., Rojanasuphot S., Ahandrik S., Viriyapongse S., et al. (1984) Risk factors in dengue shock syndrome: a prospective epidemiologic study in Rayong, Thailand. Am J Epidemiol. 120 (5): 653–669. https://academic.oup.com/aje/article-abstract/120/5/653/90744?

[40] Screaton G., Mongkolsapaya J., Yacoub S., et al. (2015) New insights into the immunopathology and control of dengue virus infection. Nat Rev Immunol. 15: 745–759. 10.1038/nri3916

[41] Sebayang A.A., Fahlena H., Anam V, Knopoff D, Stollenwerk N, Aguiar M, Soewono E. (2021) Modeling Dengue Immune Responses Mediated by Antibodies: A Qualitative Study. Biology (Basel). 10(9):941. 10.3390/biology10090941.

[42] Sierra B., Perez A.B., Vogt K., Garcia G., et al. (2010) Secondary heterologous dengue infection risk: disequilibrium between immune regulation and inflammation? Cell Immunol. 262 (2):134–140. https://www.sciencedirect.com/science/article/abs/pii/S0008874910000353

[43] Sierra B., García G., Pérez A.B., Morier L., et al. (2002) Long-term memory cellular immune response to dengue virus after a natural primary infection. International Journal of Infectious Diseases. 6(2):125–128. 10.1016/S1201-9712(02)90073-1

[44] Shaw, L.B., Billings, L., Schwartz, I.B. (2007) Using dimension reduction to improve outbreak predictability of multistrain diseases. J. Math. Biol. 55, 1–19. 10.1007/s00285-007-0074-x

[45] Shih H., Wang Y., Wang Y.P., Chi Y., Chien Y.W. (2024) Risk of severe dengue during secondary infection: A population-based cohort study in Taiwan. Journal of Microbiology, Immunology and Infection. 57(5):730–738. 10.1016/j.jmii.2024.07.004

[46] St. John A.L., Rathore A.P.S. (2019) Adaptive immune responses to primary and secondary dengue virus infections. Nat Rev Immunol. 19: 218—230. 10.1038/s41577-019-0123-x

[47] Steindorf V., Oliva S., Stollenwerk N., Aguiar M. (2024) Symmetry in a multi-strain epidemiological model with distributed delay as a general cross-protection period and disease enhancement factor. Communications in Nonlinear Science and Numerical Simulation. 128: 107663. 10.1016/j.cnsns.2023.107663

[48] Steindorf V., Oliva S., Wu J. (2022) Cross immunity protection and antibody-dependent enhancement in a distributed delay dynamic model[J]. Mathematical Biosciences and Engineering. 19(3): 2950–2984. 10.3934/mbe.2022136

[49] Steindorf V., Srivastav A.K., Stollenwerk N., et al. (2022) Modeling secondary infections with temporary immunity and disease enhancement factor: Mechanisms for complex dynamics in simple epidemiological models. Chaos, Solitons & Fractals. 164: 112709. 10.1016/j.chaos.2022.112709

[50] Steindorf V., Srivastav A.K., Stollenwerk N., et al. (2024) Beyond the biting - limited impact of explicit mosquito dynamics in dengue models. BMC Infect Dis. 24:1090. 10.1186/s12879-024-09995-6

[51] Stollenwerk N., Mateus L., Steindorf V., et al. (2025) Evaluating the risk of mosquito-borne diseases in non-endemic regions: A dynamic modeling approach. Mathematics and Computers in Simulation. 238: 1–24. 10.1016/j.matcom.2025.04.026

[52] Taghikhani R., Gumel A.B. (2018) Mathematics of dengue transmission dynamics: Roles of vector vertical transmission and temperature fluctuations. Infectious Disease Modelling. 3:266–292. 10.1016/j.idm.2018.09.003.

[53] en Bosch Q.A., Singh B.K., Hassan M.R.A., Chadee D.D., Michael E. (2016) The Role of Serotype Interactions and Seasonality in Dengue Model Selection and Control: Insights from a Pattern Matching Approach. PLOS Neglected Tropical Diseases 10(5): e0004680. 10.1371/journal.pntd.0004680

[54] van den Driessche P., Watmough J. (2008) Further notes on the basic reproduction number. In F. Brauer, P. van den Driessche, and J. Wu, editors. Mathematical Epidemiology. pp 159–178. Springer Berlin Heidelberg, Berlin, Heidelberg.

[55] Waggoner J.J., Balmaseda A., Gresh L., Sahoo M.K., Montoya M., et al. (2016) Homotypic Dengue Virus Reinfections in Nicaraguan Children. The Journal of Infectious Diseases. 214(7): 986–993. 10.1093/infdis/jiw099

[56] Woodall H., Adams B. (2014) Partial cross-enhancement in models for dengue epidemiology. Journal of Theoretical Biology. 351:67–73. 10.1016/j.jtbi.2014.02.016

[57] Zompi S., Santich B.H., Beatty P.R., Harris E. (2012) Protection from secondary dengue virus infection in a mouse model reveals the role of serotype cross-reactive B and T cells. J Immunol. 188(1):404–16. 10.4049/jimmunol.1102124

